# Sexual risk-taking behavior amongst emerging adults in Africa: a systematic review and meta-analysis

**DOI:** 10.1101/2022.09.13.22279893

**Authors:** Stevenson K. Chea, Vincent A. Kagonya, Osman Abdullahi, Amina A. Abubakar, Souheila Abbeddou, Kristien Michielsen, Amin S. Hassan

**Affiliations:** Department of Nursing Sciences, School of Health and Human Sciences, Pwani University, Kilifi, Kenya; International Centre for Reproductive Health, Department of Public Health and Primary Care, Faculty of Medicine and Health Sciences, Ghent University, Ghent, Belgium; Public Health Nutrition Unit, Department of Public Health and Primary Care, Faculty of Medicine and Health Sciences, Ghent University, Ghent, Belgium; Centre for Geographic Medicine Research, Kenya Medical Research Institute, Kilifi, Kenya; Department of Public Health, School of Health and Human Sciences, Pwani University, Kilifi, Kenya; Department of Psychiatry, University of Oxford, Oxford, United Kingdom; Institute for Human Development, Aga Khan University, Nairobi, Kenya

**Keywords:** Sexual risk-taking, emerging adults

## Abstract

**Background:** Incidence of HIV and other sexually transmitted infections (STIs) among emerging adults (EmA) is high in some African settings. Estimates on sexual risk-taking behavior (SRTB) among EmA is varied in literature, which presents a challenge when designing targeted interventions. We aimed to review and summarize literature on prevalence and risk factors of SRTB among EmA in Africa.

**Methods:** A search for studies published in PubMed, Embase and Psych Info involving EmA (18 – 25 years), conducted in Africa and reporting one or more SRTB was done. Pooled prevalence estimates were summarized using forest plots. Heterogeneity in SRTB was explored by sex, geographic region, year of publication and outcome definition. Risk factors were synthesized using a modified socio-ecological model.

**Results:** Overall, 117 studies were analyzed. Non-condom use had the highest pooled prevalence (46.0% [95% CI: 14.0-51.0]), followed by study-defined SRTB (37.0% [95% CI: 23.0 -51.0]), concurrency (35.0% [95% CI: 19.0-53.0]), multiple sex partnerships (30.0% [95% CI: 24.0-37.0]), younger age at sexual debut (25.0% [95% CI: 19.0-31.0]), age disparate relationships (24.0% [95% CI: 17.0-32.0]) and transactional sex (17.0% [95% CI: 11.0-24.0]). In four of the seven outcomes, heterogeneity was partially explained by sex, with female participants having higher pooled prevalence estimates compared to their male counterparts. In four of the seven outcomes, alcohol/drug use was the most common risk factor.

**Conclusions:** SRTB was common among EmA and differentially higher in emerging female adults. Non-condom use had the highest pooled prevalence, which may contribute to the propagation of HIV and other STIs in this population. Interventions targeting emerging female adults and alcohol/drug use may reduce SRTB, which may in-turn mitigate propagation of HIV and other STIs among EmA in Africa.

## Background

HIV-1 transmission in the general population in sub-Saharan Africa (sSA) is estimated to have declined from 1.4 million in 2010 to 800,000 in 2019 [1]. However, new HIV-1 infections among young adults continue to raise concerns as they are estimated to contribute 31% of new infections in sSA in 2019 [1]. Earlier studies from Mozambique, Uganda, Tanzania and Zimbabwe indicate that sexual risk-taking behavior (SRTB) is highly prevalent among EmA [2-6]. Literature suggests a positive linear association between SRTB and STIs, including HIV [7], which may partially explain the high rates of STIs, including HIV-1 infection among EmA in Africa.

Emerging adulthood is a relatively new concept of development for the period from late teens through the early twenties, with a focus on ages 18-25 years [8]. During this time, experimentation that began in adolescence intensifies [9]. Emerging adulthood is a distinct period demographically [10], subjectively and in terms of identity explorations [9]. Demographically, the transition from teenage hood into emerging adulthood is characterized with a move from parents’ houses to a semi-autonomous life [11]. Subjectively, EmA feel they have left adolescence but have not yet completely transitioned into adulthood [12]. Lastly, emerging adulthood is the period of life that offers most opportunity for identity explorations in the areas of work and relationships [13]. Identity formation involves trying out various life possibilities and gradually moving towards making enduring decisions and commitments. Romantic relationships at this age often include sexual intercourse [14], which likely exposes EmA to SRTB.

Another notable characteristic in emerging adulthood is SRTB. Evidence from the USA suggests that the prevalence of SRTB was higher during emerging adulthood (31.8%) compared to adolescence (21.6%) [15]. Literature on SRTB amongst EmA suggests that outcomes remain varied in estimates. For example, a recent systematic review has shown that SRTB among youth in Africa range from 3% in Zimbabwe to 89% in Uganda [16]. The review however did not focus on EmA only as it included studies that recruited HIV-1 infected individuals aged 10-24 years. In this review, the most common SRTB was non-condom use at 88.7% [16]. Another systematic review mainly focused on adolescents and reported prevalence of non-condom use ranging from 35-55% [17]. Importantly and in both studies, the potential explanation for the wide variation in SRTB prevalence estimates was not explored. Variations in SRTB estimates present a challenge in the design and implementation of effective and targeted interventions. We aimed to systematically review and summarize comparable literature on prevalence and risk factors of SRTB among EmA in Africa.

## Methods

### Search strategy

Literature published on PubMed, Embase, and Psych Info up to April 2020 was searched. The search was done using relevant key concepts and search terms based on the Population, Exposure, Comparison, Outcome and Setting (PECOS) approach (File S1). To identify articles that may have been missed in the search, snowballing was used to search reference lists of all eligible articles. The preferred reporting items for systematic reviews and meta-analyses (PRISMA) guidelines were used in reporting the methods and findings [18, 19]. The protocol for this systematic review was registered on PROSPERO (registration number CRD42020150075).

### Eligibility

Eligibility criteria for inclusion of studies in the review were: (i) participants 18-25 years old (or mean and/or median age within 18-25 years, or outcome data reported in an eligible sub-population), (ii) quantified any SRTB assessed as a primary or secondary outcome, (iii) conducted in Africa, (iv) observational studies, and (v) published in English. Two investigators (SC and VAK) screened all the articles by title/abstract then by full text for eligibility. Disagreements on eligibility were settled through consultation and consensus building.

### Outcome definition

The primary outcome was SRTB. Studies reporting one or more of six different variants or sub-variants of SRTB including: non-condom use, multiple sex partnerships, transactional sex, younger age at sexual debut, concurrency and age disparate relationships were assessed. In cases where studies reported two or more SRTBs combined as one outcome, we reported this separately as “study–defined SRTB”. Thus, and all-together, seven SRTB outcomes were assessed.

### Data extraction

Eligible articles were reviewed and relevant data extracted. These included author details, year of publication, sample size, study design, study setting, participants’ sex and age. Outcome-specific data included definition of the outcome, prevalence estimate, all the risk factors that were assessed and risk factors reported to be significantly associated with the outcome. In studies that reported different variants of an outcome, prevalence estimates for both the outcome and its variants were extracted only if the outcome definitions differed based on the recall period (e.g. main outcome [non-condom use in the past one year]; variant [non-condom use in the past 6 months]). Where the recall period was similar or not specified (e.g. main outcome [non-condom use with casual partner]; variant [non-condom use with regular partner]), only the prevalence estimate of the main outcome, as defined in the study objectives, was extracted. In studies where outcome data were stratified by sex, estimates were collapsed to obtain an overall estimate. Similarly, for such studies, factors reported to be significantly associated with reported outcomes were extracted only if they were significant in both sexes with a similar direction of association. This approach was taken to ensure consistency in reporting of risk factors. Data abstraction was independently conducted by two investigators (SC and VK) and a comparison of results done. Disagreements were settled through consultations and consensus building.

### Quality assessment

The quality assessment tool for systematic reviews of observational studies (QATSO) was used to appraise the quality of eligible studies. The QATSO tool has been shown to be practical, simple, and easy when reviewing observational studies in the context of health risk outcomes and has been shown to have good inter-rater reliability [20]. During data extraction, each study that met the inclusion criteria was reviewed against the five risk-of-bias items that check on external validity (1 item), bias (1 item), reporting (2 items), and confounding (1 item). Items were coded and scored as 1 if the condition was met and 0 if not. The summed score was then divided by the total number of all applicable items. Next, the resultant score was converted to a percentage that guided the generation of a ‘traffic light’ rating of a study as “Poor” (less than 33%), “Satisfactory” (33-66%), or “Good” (greater than 66%). All eligible studies regardless of quality score were included in analysis.

### Data analysis

Prevalence estimates from the seven pre-determined SRTB outcomes were summarized and presented using forest plots. Heterogeneity in the prevalence estimates was further explored by sex, geographical region where studies were conducted, year of publication, and outcome definition. Weighted individual proportions and their variances were first computed. Since proportional data from observational studies is often skewed [21], observed proportions from individual studies were then transformed using the double arcsine transformation [22]. Secondly, the transformed proportions and their sampling variances were pooled to generate summary proportions. To consider within and between study variances and to allow generalizability of the findings, random effect models were used to calculate the summary proportions [23]. Random effects modeling was estimated using the DerSimonian and Laird method. In this method, the summary proportion was estimated as the weighted average of the observed proportion of individual studies where the weighting for each study is the inverse of the total variance of a study [24]. In this case, total variance was the sum of within study and between study variance. Finally, the transformed proportions were converted back and pooled prevalence estimates reported. Analysis was performed in Stata 15.0 (StataCorp.2017. Stata Statistical Software: Release 15. College Station, TX: StataCorp LLC. 2019).

Risk factors of each SRTB outcome were synthesized, summarized, and grouped into five thematic categories based on a modified socio-ecological model [25, 26]: (i) Socio-demographic, (ii) relationship and behavioral, (iii) knowledge, attitude and beliefs, (iv) family and community and (v) mental and physical health factors. For studies that reported variants of the outcomes, only risk factors from the main SRTB outcome were included in the synthesis.

## Results

### Characteristics of eligible studies

From the three databases, we obtained a combined 4,280 hits (PubMed [n=720], Psych info [n=1,323] and Embase [n=2,237]) of which 1,139 were duplicates. A further 3,024 articles were also excluded. The remaining 117 articles met all eligibility criteria and were included in the analysis (Fig 1). Overall, eligible studies comprised 163,251 EmA, with sample sizes ranging from 50 to 27,757 participants. In summary, majority of eligible studies were published after 2010 (n=89 [76.1%]), used a cross sectional study design (n=109 [93.2%]), had a sample size of less than 1000 participants (n= 75 [64.1%]), recruited both male and female participants (n=81 [69.2%]), and were from the Eastern Africa region (n=57 [48.7%]) (Table 1). The most reported SRTB outcome was non-condom use (n=93 [79.5%]). Overall, 36 (30.7%) studies reported SRTB outcomes and their variants. Based on the QATSO, a majority of studies had a good quality score (n=91 [77.8%]) (Table 2).

**Fig 1:**
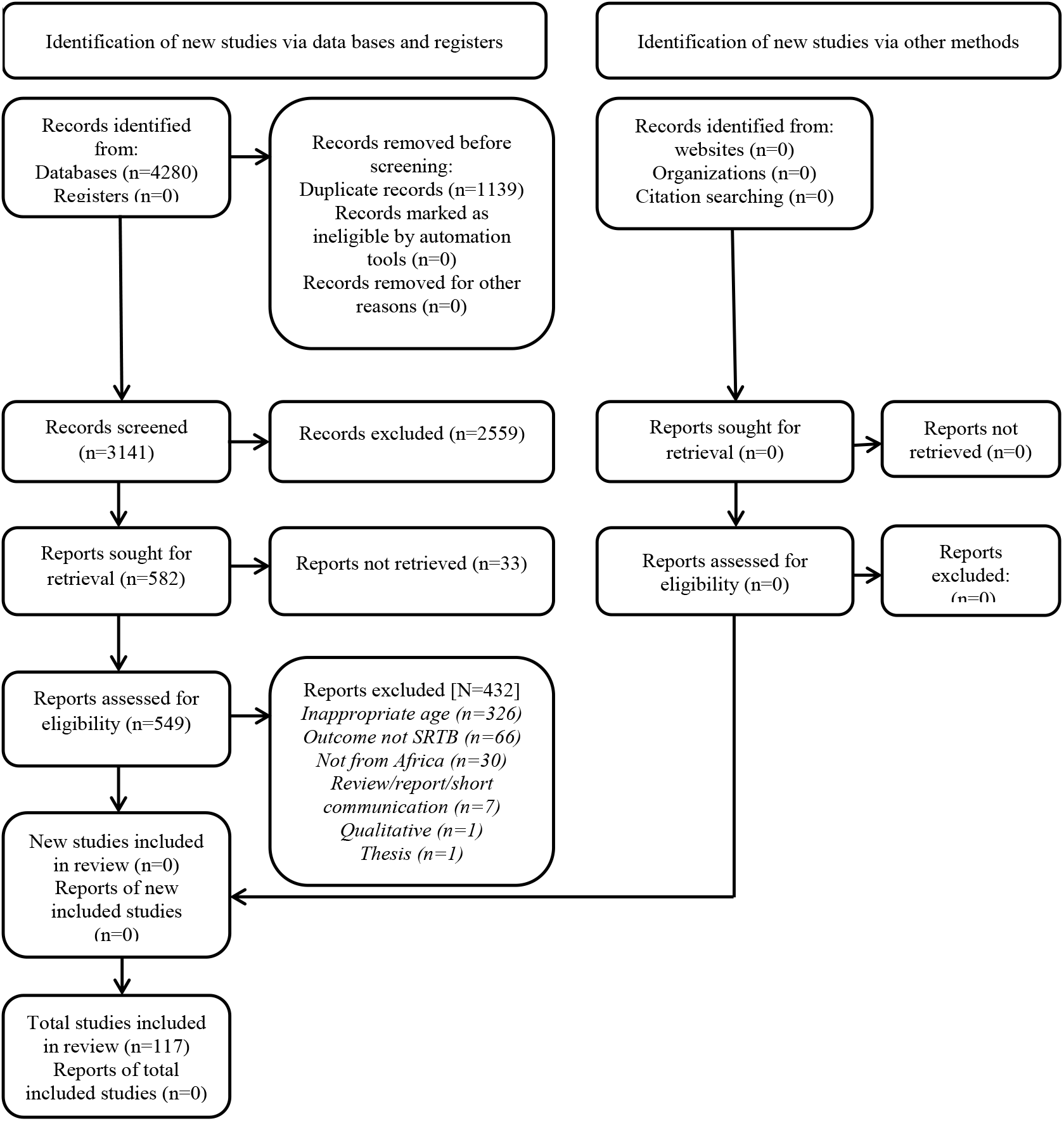
PRISMA flow chart illustrating how number of articles included in analysis was arrived at.

**Table 1:**
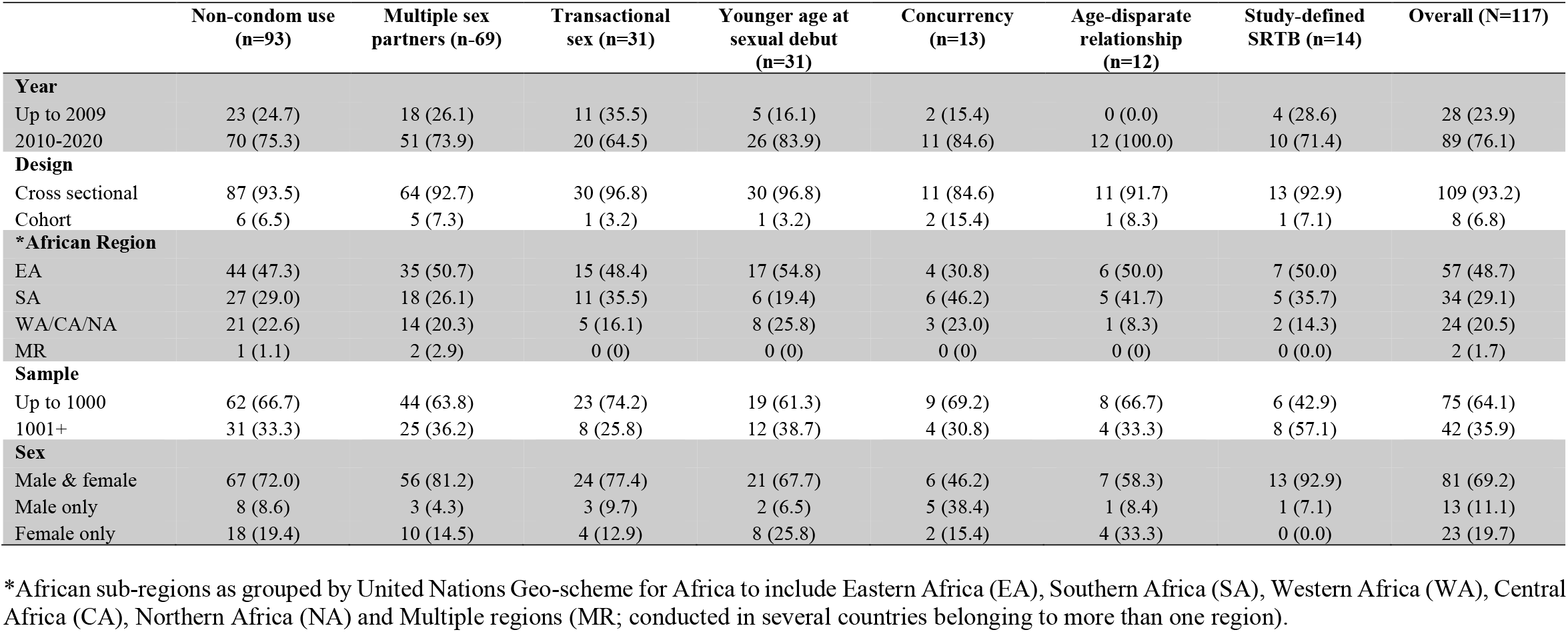
Description of studies included in the analysis by year of publication, study design, African region in which individual studies were conducted, sample size and sex of study participants (n=117)

|  | Non-condom use<br>(n=93) | Multiple sex<br>partners (n=69) | Transactional<br>sex (n=31) | Younger age at<br>sexual debut<br>(n=31) | Concurrency<br>(n=13) | Age-disparate<br>relationship<br>(n=12) | Study-defined<br>SRTB (n=14) | Overall (N=117) |
| --- | --- | --- | --- | --- | --- | --- | --- | --- |
| **Year** |  |  |  |  |  |  |  |  |
| Up to 2009 | 23 (24.7) | 18 (26.1) | 11 (35.5) | 5 (16.1) | 2 (15.4) | 0 (0.0) | 4 (28.6) | 28 (23.9) |
| 2010-2020 | 70 (75.3) | 51 (73.9) | 20 (64.5) | 26 (83.9) | 11 (84.6) | 12 (100.0) | 10 (71.4) | 89 (76.1) |
| **Design** |  |  |  |  |  |  |  |  |
| Cross sectional | 87 (93.5) | 64 (92.7) | 30 (96.8) | 30 (96.8) | 11 (84.6) | 11 (91.7) | 13 (92.9) | 109 (93.2) |
| Cohort | 6 (6.5) | 5 (7.3) | 1 (3.2) | 1 (3.2) | 2 (15.4) | 1 (8.3) | 1 (7.1) | 8 (6.8) |
| **\*African Region** |  |  |  |  |  |  |  |  |
| EA | 44 (47.3) | 35 (50.7) | 15 (48.4) | 17 (54.8) | 4 (30.8) | 6 (50.0) | 7 (50.0) | 57 (48.7) |
| SA | 27 (29.0) | 18 (26.1) | 11 (35.5) | 6 (19.4) | 6 (46.2) | 5 (41.7) | 5 (35.7) | 34 (29.1) |
| WA/CA/NA | 21 (22.6) | 14 (20.3) | 5 (16.1) | 8 (25.8) | 3 (23.0) | 1 (8.3) | 2 (14.3) | 24 (20.5) |
| MR | 1 (1.1) | 2 (2.9) | 0 (0) | 0 (0) | 0 (0) | 0 (0) | 0 (0.0) | 2 (1.7) |
| **Sample** |  |  |  |  |  |  |  |  |
| Up to 1000 | 62 (66.7) | 44 (63.8) | 23 (74.2) | 19 (61.3) | 9 (69.2) | 8 (66.7) | 6 (42.9) | 75 (64.1) |
| 1001+ | 31 (33.3) | 25 (36.2) | 8 (25.8) | 12 (38.7) | 4 (30.8) | 4 (33.3) | 8 (57.1) | 42 (35.9) |
| **Sex** |  |  |  |  |  |  |  |  |
| Male & female | 67 (72.0) | 56 (81.2) | 24 (77.4) | 21 (67.7) | 6 (46.2) | 7 (58.3) | 13 (92.9) | 81 (69.2) |
| Male only | 8 (8.6) | 3 (4.3) | 3 (9.7) | 2 (6.5) | 5 (38.4) | 1 (8.4) | 1 (7.1) | 13 (11.1) |
| Female only | 18 (19.4) | 10 (14.5) | 4 (12.9) | 8 (25.8) | 2 (15.4) | 4 (33.3) | 0 (0.0) | 23 (19.7) |

\*African sub-regions as grouped by United Nations Geo-scheme for Africa to include Eastern Africa (EA), Southern Africa (SA), Western Africa (WA), Central Africa (CA), Northern Africa (NA) and Multiple regions (MR; conducted in several countries belonging to more than one region).

**Table 2:** Distribution of studies included in analysis by their quality as assessed using the QUATSO tool (n=117)

| Quality score (%) | Quality assessment | Number of studies n (%) |
| --- | --- | --- |
| <33 | Poor | 6 (5.1) |
| 33-66 | Satisfactory | 20 (17.1) |
| >66 | Good | 91 (77.8) |

### Pooled prevalence and risk factors of sexual risk-taking behavior

#### i) Non-condom use

Overall, 93 (79.5%) studies reported non-condom use, thirty of which comprised a cumulative forty-four sub-variants of the outcome yielding 137 records. Non-condom use at last sexual encounter was the most used definition (n=42 [30.7%]) (S1 Table). The pooled prevalence of non-condom use was 46.0% (95% CI: 41.0-51.0) (A in S1 Fig). There was significant heterogeneity between studies (I^2^=99.7%, p<0.01), with female-only studies and both-sex studies (compared to male-only studies, p=0.04), studies from the Eastern Africa region (compared to other regions, p<0.01) and studies published before 2010 (compared to those published after 2010, p=0.01) having significantly higher pooled estimates (Table 3 and B, C, D and E in S1 Fig).

**Table 3:** Overall pooled prevalence estimates of the seven SRTB outcomes assessed in the current review as well as pooled estimates by sex of study participants, African region where studies were conducted, year of publication and definition of individual SRTB outcomes in primary studies; and outstanding/frequently reported risk factors and their categories for all outcomes and their variants in the included studies (n=117)

|  | Non-Condom use (n=93) | Multiple sex partners (n=69) | Transactional sex (n=31) | Younger age at sexual debut (n=31) | Concurrency (n=13) | Age-disparate relationship (n=12) | Study defined SRTB (n=14) |
| --- | --- | --- | --- | --- | --- | --- | --- |
| **Total records** | 137 | 74 | 32 | 31 | 13 | 12 | 14 |
| Pooled prevalence, % (95% CI) | 46 (41-51) | 30 (24-37) | 17 (11-24) | 25 (19-31) | 35 (19-53) | 24 (17-32) | 37 (23-51) |
| Total heterogeneity, % (p value) | 99.7 (p<0.01) | 99.7 (p<0.01) | 99.2 (p<0.01) | 99.1 (p<0.01) | 99.6 (p<0.01) | 98.4 (p<0.01) | 99.6 (p<0.01) |
| **Sub-group analysis by sex** |  |  |  |  |  |  |  |
| *Both male & female* |  |  |  |  |  |  |  |
| Pooled prevalence (%); 95% CI | 47 (41-53) | 30 (23-37) | 13 (7-20) | 23 (17-29) | 24 (17-32) | 23 (15-32) | 39 (25-53) |
| Heterogeneity (%); p value | 99.7 (p<0.01) | 99.8 (p<0.01) | 99.1 (p<0.01) | 98.9 (p<0.01) | 95.7 (p<0.01) | 98.1 (p<0.01) | 99.5 (p<0.01) |
| *Male* |  |  |  |  |  |  |  |
| Pooled prevalence (%); 95% CI | 29 (18-42) | 40 (3-85) | 28 (13-45) | 29 (26-33) | 57 (18-91) | 3 (1-6) *** | 13 (11-14) *** |
| Heterogeneity (%); p value | 99.2 (p<0.01) | * | * | * | 99.8 (p<0.01) | * | * |
| *Female* |  |  |  |  |  |  |  |
| Pooled prevalence (%); 95% CI | 47 (39-55) | 29 (18-41) | 32 (16-52) | 29 (11-51) | 16 (12-20) | 34 (24-45) |  |
| Heterogeneity (%); p value | 99.2 (p<0.01) | 98.9 (p<0.01) | 99.0 (p<0.01) | 99.4 (p<0.01) | * | 97.1 (p<0.01) |  |
| Heterogeneity between groups (p value) | p=0.04 | p=0.91 | p=0.04 | p=0.21 | p=0.02 | p<0.01 | p<0.01 |
| **Sub-group analysis by region where study was conducted** |  |  |  |  |  |  |  |
| *Eastern Africa* |  |  |  |  |  |  |  |
| Pooled prevalence (%); 95% CI | 49 (43-55) | 28 (23-34) | 14 (8-22) | 20 (15-25) | 49 (14-86) | 31 (24-38) | 30 (18-43) |
| Heterogeneity (%); p value | 99.4 (p<0.01) | 98.8 (p<0.01) | 98.6 (p<0.01) | 97.4 (p<0.01) | 99.8 (p<0.01) | 95.8 (p<0.01) | 99.1 (p<0.01) |
| *Western/Central/Northern Africa* |  |  |  |  |  |  |  |
| Pooled prevalence (%); 95% CI | 48 (37-59) | 33 (15-54) | 19 (13-25) | 39 (18-62) | 21 (12-31) | 29 (25-34) *** | 39 (37-42) |
| Heterogeneity (%); p value | 99.7 (p<0.01) | 99.8 (p<0.01) | 91.8 (p<0.01) | 99.5 (p<0.01) | * | * | * |
| *Southern Africa* |  |  |  |  |  |  |  |
| Pooled prevalence (%); 95% CI | 36 (27-44) | 33 (24-43) | 20 (8-36) | 24 (14-34) | 33 (9-64) | 16 (7-28) | 47 (17-78) |
| Heterogeneity (%); p value | 99.4 (p<0.01) | 99.3 (p<0.01) | 99.6 (p<0.01) | 99.2 (p<0.01) | 99.7 (p<0.01) | 98.6 (p<0.01) | 99.8 (p<0.01) |
| *Multiple regions* |  |  |  |  |  |  |  |
| Pooled prevalence (%); 95% CI | 82 (81-83) *** | 17 (16-17) |  |  |  |  |  |
| Heterogeneity (%); p value | * | * |  |  |  |  |  |
| Heterogeneity between groups (p value) | p<0.01 | p<0.01 | p=0.60 | p=0.19 | p=0.29 | p=0.10 | p=0.37 |
| **Sub-group analysis by year of publication** |  |  |  |  |  |  |  |
| *Up to 2009* |  |  |  |  |  |  |  |
| Pooled prevalence (%); 95% CI | 57 (48-66) | 27 (19-36) | 16 (6-29) | 17 (13-21) | 36 (34-38) | ** | 33 (5-70) |
| Heterogeneity (%); p value | 99.6 (p<0.01) | 99.2 (p<0.01) | 99.5 (p<0.01) | 94.8 (p<0.01) | * | ** | 99.8 (p<0.01) |
| *2010-2020* |  |  |  |  |  |  |  |
| Pooled prevalence (%); 95% CI | 43 (37-49) | 31 (23-40) | 17 (10-25) | 27 (19-35) | 33 (16-53) | ** | 38 (23-55) |
| Heterogeneity (%); p value | 99.7 (p<0.01) | 99.8 (p<0.01) | 98.9 (p<0.01) | 99.1 (p<0.01) | 99.5 (p<0.01) | ** | 99.5 (p<0.01) |
| Heterogeneity between groups (p value) | p=0.01 | p=0.46 | p=0.90 | p=0.02 | p=0.81 | ** | p=0.79 |
| **Sub-group analysis by outcome definition** |  |  |  |  |  |  |  |
| *12 months or less* |  |  |  |  |  |  |  |
| Pooled prevalence (%); 95% CI | 43 (34-53) | 28 (24-32) | 20 (9-36) |  | 47 (22-73) |  | 28 (11-49) |
| Heterogeneity (%); p value | 99.5 (p<0.01) | 99.1 (p<0.01) | 98.9 (p<0.01) |  | 99.4 (p<0.01) |  | 99.7 (p<0.01) |
| *Unspecified recall period* |  |  |  |  |  |  |  |
| Pooled prevalence (%); 95% CI | 47 (36-58) | 42 (25-59) | 16 (8-25) |  | 22 (6-44) |  | 65 (40-86) |
| Heterogeneity (%); p value | 99.7 (p<0.01) | 99.5 (p<0.01) | 99.3 (p<0.01) |  | 99.7 (p<0.01) |  | * |
| *Lifetime/ever* |  |  |  |  |  |  |  |
| Pooled prevalence (%); 95% CI |  | 30 (20-41) | 13 (4-27) |  |  |  |  |
| Heterogeneity (%); p value |  | 98.4 (p<0.01) | 98.4 (p<0.01) |  |  |  |  |
| *Last sex* |  |  |  |  |  |  |  |
| Pooled prevalence (%); 95% CI | 45 (36-53) |  |  |  |  |  |  |
| Heterogeneity (%); p value | 99.6 (p<0.01) |  |  |  |  |  |  |
| *Never/first sex* |  |  |  |  |  |  |  |
| Pooled prevalence (%); 95% CI | 50 (39-61) |  |  |  |  |  |  |
| Heterogeneity (%); p value | 99.5 (p<0.01) |  |  |  |  |  |  |
| *First sex before age 18 years* |  |  |  |  |  |  |  |
| Pooled prevalence (%); 95% CI |  |  |  | 24 (19-29) |  |  |  |
| Heterogeneity (%); p value |  |  |  | 98.7 (p<0.01) |  |  |  |
| *First sex at age 18-20 years* |  |  |  |  |  |  |  |
| Pooled prevalence (%); 95% CI |  |  |  | 32 (6-68) |  |  |  |
| Heterogeneity (%); p value |  |  |  | 99.6 (p<0.01) |  |  |  |
| *Age difference 4 years or more* |  |  |  |  |  |  |  |
| Pooled prevalence (%); 95% CI |  |  |  |  |  | 26 (19-33) |  |
| Heterogeneity (%); p value |  |  |  |  |  | 97.0 (p<0.01) |  |
| *Age difference unspecified* |  |  |  |  |  |  |  |
| Pooled prevalence (%); 95% CI |  |  |  |  |  | 12 (11-13) |  |
| Heterogeneity (%); p value |  |  |  |  |  | * |  |
| *Lifetime/ever/last sex* |  |  |  |  |  |  |  |
| Pooled prevalence (%); 95% CI |  |  |  |  |  |  | 33 (25-41) |
| Heterogeneity (%); p value |  |  |  |  |  |  | * |
| Heterogeneity between groups (p value) | p=0.81 | p=0.29 | p=0.75 | p=0.60 | p=0.14 | p<0.01 | p=0.04 |

\*Three or less studies in sub-group hence no heterogeneity estimates
\*\*All studies fall in one category hence no sub-groups
\*\*\*Prevalence estimate is for one study only since that is the only study in the sub-group

Of the 93 studies, 57 (61.2%) assessed risk factors for non-condom use (S2 Table). Of these, 42 reported at least one significant factor associated with non-condom use. Alcohol/illicit drug use was the most common risk factor for non-condom use (n=13 studies) (Table 4).

**Table 4:**
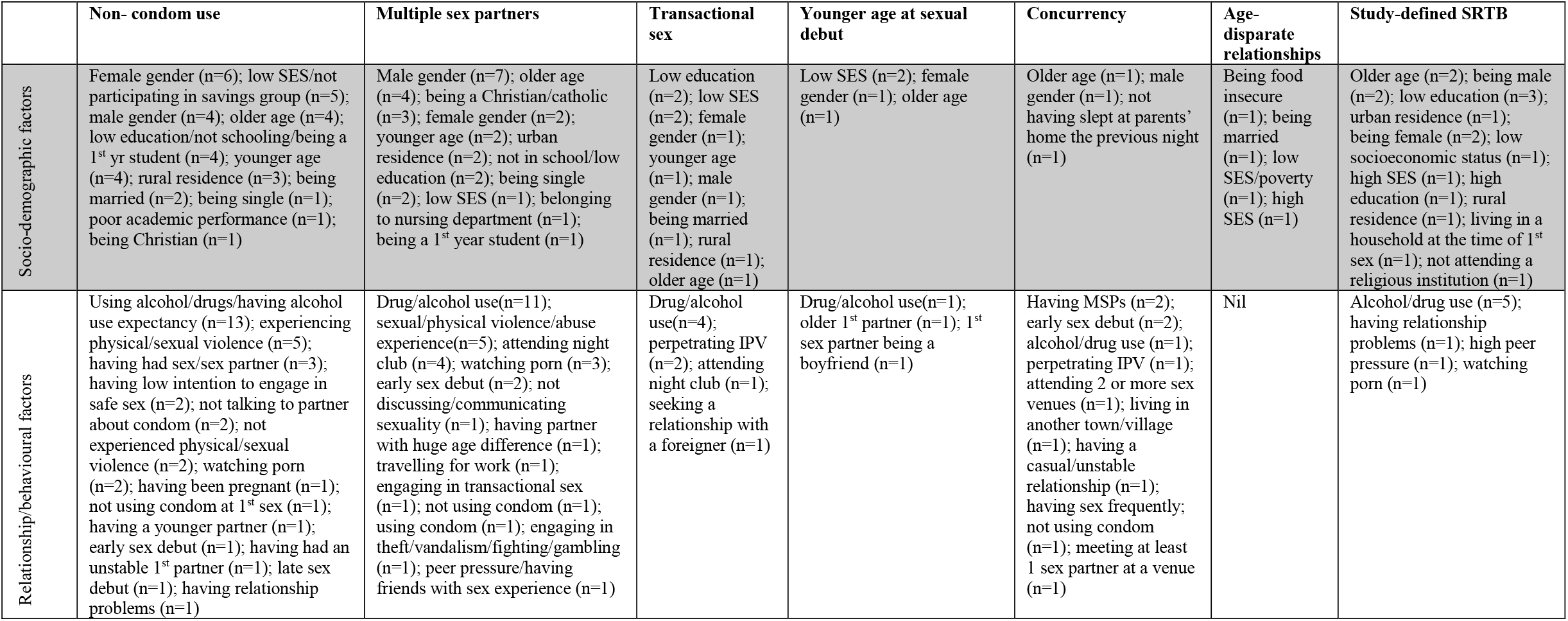
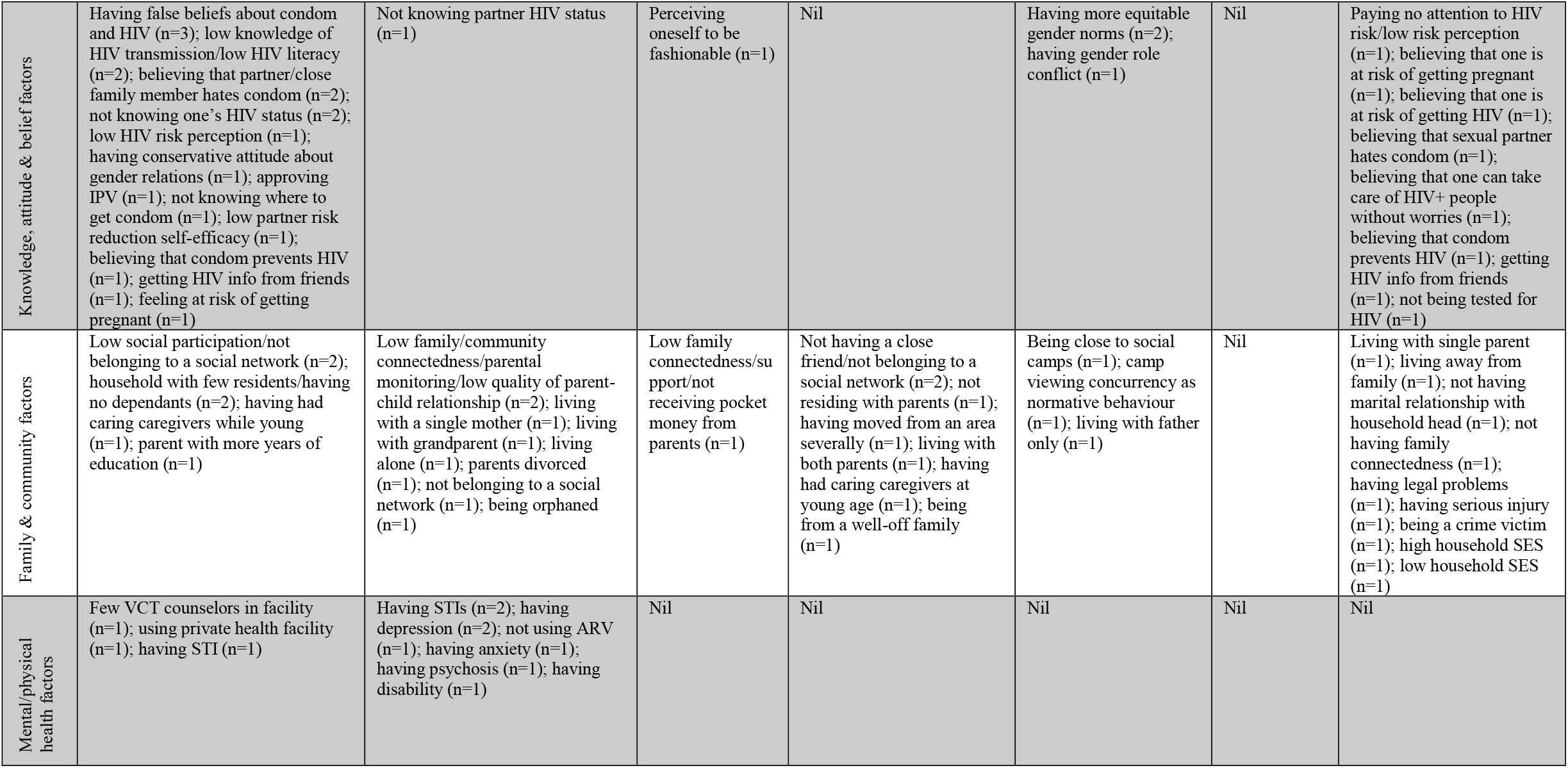
Summary of factors reported to be significantly associated with SRTB outcomes in studies included in analysis grouped in five thematic areas derived from a modified socio-ecological model.

#### ii) Multiple sex partnerships (MSP)

Overall, 69 (59.0%) studies reported MSP, five of which included sub-variants of the outcome yielding 74 records. The majority of studies (n=57 [77.0%]) defined the outcome as having MSP in the last 12 months or less (S1 Table). The pooled prevalence of MSP was 30.0% (95% CI: 24.0-37.0) (A in S2 Fig). There was significant heterogeneity between studies (I^2^=99.7%, p<0.01), with studies from both the Western Africa/Central Africa/Northern Africa (WA/CA/NA) and Southern Africa (SA) regions having equal and significantly higher pooled prevalence estimates compared to the other regions (p=0.01) (Table 3 and B, C, D and E in S2 Fig).

Of the 69 studies, 35 (50.7%) assessed risk factors for MSP (S2 Table). Of these, 27 reported at least one significant factor associated with MSP. Alcohol/illicit drug use was the most common risk factor for MSP (n=11 studies) (Table 4).

#### iii) Transactional sex (TS)

Overall, 31 (26.5%) studies reported TS, one of which included a sub-variant of the outcome yielding 32 records. The majority of studies (n=18 [56.3%]) defined TS without including a recall period (S1 Table). The pooled prevalence of TS was 17.0% (95% CI: 11.0-24.0) (A in S3 Fig). There was significant heterogeneity between studies (I^2^=99.2%, p<0.01), with female-only studies (compared to male-only studies and both-sex studies, p=0.04) having significantly higher prevalence estimates (Table 3 and B, C, D and E in S3 Fig).

Of the 31 studies, 12 (38.7%) assessed risk factors for TS (S2 Table). Of these, 11 reported at least one significant factor associated with TS. Alcohol/illicit drug use was the most common risk factor for TS (n=4 studies) (Table 4).

#### iv) Younger age at sexual debut (YASD)

Overall, 31 (26.5%) studies reported YASD. None included a sub-variant of the outcome. The majority of studies (n=26 [83.9%]) defined YASD as having first sex before the age of 18 years (S1 Table). The pooled prevalence of YASD was 25.0% (95% CI: 19.0-31.0) (A in S4 Fig). There was significant heterogeneity between studies (I^2^=99.1%, p<0.01), with studies published after 2010 having higher pooled prevalence estimate, compared to those published prior 2010 (p=0.02) (Table 3 and B, C, D and E in S4 Fig).

Of the 31 studies, 10 (32.2%) assessed risk factors for YASD (S2 Table). Of these, 6 reported at least one significant factor associated with YASD. Low socioeconomic status and not belonging to a social network were the two outstanding risk factors for YASD (n=2 studies each) (Table 4).

#### v) Concurrency

Overall, 13 (11.1%) studies reported concurrency. None included a sub-variant of the outcome, yielding 13 records. The majority of studies defined concurrency as having concurrent sexual partners in the last 12 months or less (n=7 [53.9%]) (S1 Table). The pooled prevalence of concurrency was 35.0% (95% CI: 19.0-53.0) (A in S5 Fig). There was significant heterogeneity between studies (I^2^=99.6%, p<0.01), with male-exclusive studies (compared to female-exclusive studies and both-sex studies, p<0.02) having higher pooled estimates (Table 3 and B, C, D and E in S5 Fig).

Of the 13 studies, 9 (69.2%) assessed risk factors for concurrency (S2 Table). Of these, 7 reported at least one significant factor associated with concurrency. Having multiple sex partners, younger age at sexual debut and having equitable gender norms were standing out as risk factors for concurrency, (n=2 studies each) (Table 4).

#### vi) Age-disparate relationships

Overall, 12 (10.2%) studies reported age-disparate relationships. None included a sub-variant of the outcome, yielding 12 records. The majority of studies defined age-disparate relationships as an age difference of four years or more between sexual partners (n=10 [83.3%]) (S1 Table). The pooled prevalence of age-disparate relationships was 24.0% (95% CI: 17.0-32.0) (A in S6 Fig). There was significant heterogeneity between studies (I^2^=98.4%, p<0.01), with female-exclusive studies (compared to male-exclusive studies and both-sex studies, p<0.01) and studies using definition of age difference of four or more years between sexual partners (compared to studies not specifying an age difference between sexual partners, p<0.01) having higher pooled prevalence estimates (Table 3 and B, C, D and E in S6 Fig).

Of the 12 studies, only 3 (25.0%) assessed risk factors for age-disparate relationships (S2 Table). Of these, two reported at least one significant factor associated with age-disparate relationships. Being food insecure, being married, low socio-economic status and high socio-economic status were each reported to be significantly associated with age-disparate relationships (n=1 study each).

#### vii) Study-defined SRTB

Overall, 14 (11.9%) studies reported study-defined SRTB. None included a sub-variant of the outcome, yielding 14 records. The majority of studies defined the outcome as engaging in SRTB in the last 12 months or less (n=8 [57.2%]) (S1 Table). The pooled prevalence of study-defined SRTB was 37.0% (95%CI: 23.0-51.0) (A in S7 Fig). There was significant heterogeneity between studies (I^2^ =99.6%, p<0.01]), with both-sex studies (compared to male-exclusive studies, p<0.01) and studies defining SRTB with no recall period (compared to studies defining study-defined SRTB with a recall period of 12 months or less and those defining it as study-defined SRTB in life-time, p=0.04) having higher pooled prevalence estimates (Table 3 and B, C, D and E in S7 Fig).

Of the 14 studies, 13 (92.8%) assessed risk factors for study-defined SRTB (S2 Table). Of these, 9 reported at least one significant factor associated with study-defined SRTB. Alcohol/illicit drug use was the most common risk factor for study-defined SRTB (n=5 studies) (Table 4).

## Discussion

Estimates of SRTB are varied in literature. We aimed to systematically review and summarize prevalence and risk factors of SRTB among EmA in Africa. Consistent with previous reviews from Africa, our overall findings suggest that SRTB in Africa is common, ranging from about one in every five (transactional sex) to one in every two (non-condom use) EmA engaging in some form of SRTB.

Our findings demonstrate that non-condom use was the most assessed SRTB. This is likely because it has strongly been associated with HIV and other sexually transmitted infections (1) and has been reported to be prevalent in populations. Indeed, we observed that about one in two EmA reported non-condom use. Our estimate is comparable to the 35.0-89.0% prevalence range reported in reviews of studies among adolescents and young adults from sSA [16, 17], but much higher than the 27.5% reported among adolescents from the United States [27] and the 33% reported among female undergraduate students from Vietnam [28].

Our review also observed that about one in three EmA from Africa engaged in MSP. While this may generally be considered high, the estimate is indeed comparable to other reviews summarizing literature from adolescents and young adults in Africa reporting MSP estimates ranging from 2.5-88.2% [16, 17]. Further, these are also consistent with the 26.0-31.4% estimates among adolescents and young adults from the US [29, 30], suggesting that the burden of MSP among EmA in Africa may be comparable to that from non-African settings.

We also observed that about one in six EmA from Africa reported TS. While this may be considered comparable to another review reporting a pooled prevalence of 20.1% among HIV-infected adolescents in sSA [31], the combined estimates are much lower compared to the 20.6-60.0% range reported in another review involving HIV-infected adolescents aged 10-24 years from sSA [16]. Further, a study of HIV-infected adolescents and young men who have sex with men (MSM) from the USA reported a 27.8% prevalence of TS [32] which is higher than our estimate. The distinct difference in estimates reported in the three reviews from sSA and the US study may be a reflection of the complexities involved in the definition of TS in different African contexts [16] and may warrant development and validation of a standardized and context relevant approach in the definition of TS in Africa.

Our findings also suggest that one in four EmA in Africa initiate sex at a younger age, with most studies using less than 18 years old as YASD. On the one hand, our estimate is comparatively lower compared to the 25.5-42.1% range reported from another review involving studies among HIV-infected adolescents aged 10-19 years from sSA [31]. The higher estimate reported in the latter review may be attributed to the inclusion of studies involving younger (10-19 years), which is therefore inherently more likely to bias the estimate on the higher side, compared to our inclusion of EmA aged 18-25 years. On the other hand, our estimate is comparatively higher compared to the 10% reported from another review involving studies among HIV-infected adolescents age 10-19 years from sSA [17]. The lower estimate in the latter review may be attributed to the inclusion of HIV-infected adolescents. By virtue of their HIV infection status, this population is likely to delay sexual debut, as has been reported elsewhere [33]. Further, studies from the USA report YASD estimates ranging from 16.0-28.0% [27, 34], which is consistent to our findings and suggest that YASD is common and comparable across the two regions.

We also observed that about one in three EmA in Africa engage in concurrent sexual relationships. To our knowledge, we could not find a systematic review of literature on concurrency amongst adolescents and young adults in Africa. A study from the USA among African American adolescent females aged 15-21 years reported a concurrency prevalence of 23.3% [30], which is comparatively lower compared to our estimate. Combined, these estimates may suggest that concurrency is much more common in the African setting, which may be attributed to low socio-economic status that forces EmA to have concurrent sexual partners for material gain. Indeed, our review shows that engaging in casual sex for monetary gain stood out as a risk factor for concurrency[35].

We also reported that about one in four EmA in Africa engaged in age-disparate relationships, with most studies defining this as an age difference of more than four years between sexual partners. Our estimate was comparable to the range of 4.0-65.7% prevalence reported from a review of studies among HIV-infected adolescents aged 10-24 years from Africa [16], but higher compared to the range of 10-17% prevalence reported in three studies among adolescents and young adults from the USA [36-38]. The higher prevalence of age-disparate relationships among EmA from Africa may be explained by the low socio-economic status in the African context, with adolescents and younger adults, mostly from disadvantaged backgrounds, engaging in relationships with older, more established partners for material gains, as has been reported elsewhere [39]. Indeed, and in our findings, low socio-economic status stood out as a risk factor for age-disparate relationships [3, 40].

There was significant between-studies heterogeneity in the prevalence estimates across all seven SRTB outcomes. Most heterogeneity was partially explained by variations from sex, with female participants having higher pooled prevalence in four of the seven outcomes, compared to their male counterparts. Differences in the way emerging female adults are socialized in the African context may have impacted on their increased SRTB [39, 41]. In addition, low socio-economic status, along with cultural practices like child marriage and coercion of young girls into early sexual activities with adults are common and may have led to increased SRTB among emerging female adults [39].

Further, alcohol/illicit drug use stood out as the most common risk factor in four of the seven outcomes. Alcohol/illicit drug use among EmA in African settings is common. Indeed, a review of studies among 15–24-year-olds in Eastern Africa reported 52.0% median prevalence of ever using alcohol [42], with some groups like university students reporting up to 89.0% prevalence of alcohol/illicit drug use [42]. Alcohol/substance use is known to impair inhibitory control [43] and is therefore not surprising that this leads to increased SRTB [43].

A major strength of our review is the inclusion of multiple SRTBs. However, our findings are not without limitations. First, SRTB outcomes were self-reported in most of the eligible studies, which may have led to an over- or underestimation of outcomes. However, use of Audio Computer-Assisted Interview (ACASI) for data collection in some of the studies may have reduced reporting bias [44]. Second, all eligible studies were included in the review regardless of their quality. Inclusion of low-quality studies may raise concerns on the rigor of our SRTB estimates. However, only a small proportion of the studies (5.1%) were of low quality, and this is unlikely to have had a significant impact on the overall findings. Last, the majority (93%) of studies included in the review were cross-sectional in design, which makes it impossible to determine directionality of associations for risk factors. However, this was beyond the scope of the current review. Still, the paucity of longitudinal SRTB studies warrants more rigorous longitudinal studies to better understand causal pathways and direction of association for risk factors among EmA in Africa.

## Conclusion and recommendations

Overall, our findings suggest that SRTB among EmA is common in the African setting. Non-condom use was most frequently assessed and had the highest, while transactional sex had the lowest pooled prevalence estimate. There was significant between-studies heterogeneity in the pooled prevalence estimates, with female participants tending to have higher SRTB estimates than their male counterparts. Alcohol/illicit drug use stood out as the most associated risk factor for SRTB. Interventions targeting emerging female adults’ alcohol/illicit drug use may reduce SRTB, which may in-turn mitigate propagation of HIV and other STIs amongst EmA in Africa.

## Data Availability

All data produced in the present study are available upon reasonable request to the authors

## Author contributions

Conceptualization - SC, AH

Data curation – SC, AH, VK

Formal analysis – SC, AH

Funding acquisition – AH, AA

Investigation – SC, VK

Methodology – SC, AH, KM, SA

Project administration – SC, OA

Resources – SA, KM

Supervision – KM, SA, AH, AA, OA

Validation – KM, SA, AH, AA, VK, OA

Visualization – SC, AH

Writing-original draft preparation – SC

Writing – review and editing – KM, SA, AH, AA, VK, OA

## Supporting information

**S1 File. Full search syntax**

**S2 File. PRISM 2009 checklist**

**S1 Table. Definitions of SRTB outcomes as reported in primary studies**. Table showing definitions of the seven SRTB outcomes as reported in the 117 primary studies and their variants summarized and grouped based on recall period.

**S2 Table. Factors assessed for their association with SRTB outcomes**. Table showing Summary of factors assessed for their association with individual SRTB outcomes in studies included in analysis grouped in five thematic areas derived from a modified socio-ecological model.

**S1 Fig. Forest plots showing pooled prevalence of non-condom use**. (A) Forest plot illustrating pooled prevalence of non-condom use from the 137 records (obtained from 93 studies that assessed non-condom use). (B) Forest plot illustrating pooled prevalence of non-condom use grouped by sex of the study participants. (C) Forest plot illustrating pooled prevalence of non-condom use grouped by the African region in which the primary study was conducted. (D) Forest plot illustrating pooled prevalence of non-condom use grouped by the year of publication of primary studies. (E) Forest plot illustrating pooled prevalence of non-condom use grouped by the definition of non-condom use in primary studies.

**S2 Fig. Forest plots showing pooled prevalence of multiple sex partners**. (A) Forest plot illustrating pooled prevalence of multiple sex partners (MSP) from the 74 records (obtained from 69 studies that assessed MSP). (B) Forest plot illustrating pooled prevalence of MSP from the 74 records grouped by sex of the study participants. (C) Forest plot illustrating pooled prevalence of MSP grouped by the African region in which the primary study was conducted. (D) Forest plot illustrating pooled prevalence of MSP grouped by the year of publication of primary studies. (E) Forest plot illustrating pooled prevalence of MSP grouped by the definition of MSP in primary studies.

**S3 Fig. Forest plots showing pooled prevalence of transactional sex**. (A) Forest plot illustrating pooled prevalence of transactional sex (TS) from the 32 records (obtained from 31 studies that assessed TS). (B) Forest plot illustrating pooled prevalence of TS from the 32 records grouped by sex of the study participants. (C) Forest plot illustrating pooled prevalence of TS grouped by the African region in which the primary study was conducted. (D) Forest plot illustrating pooled prevalence of TS by year of publication of primary studies. (E) Forest plot illustrating pooled prevalence of TS grouped by the definition of TS in primary studies.

**S4 Fig. Forest plots showing pooled prevalence of younger age at sexual debut**. (A) Forest plot illustrating pooled prevalence of younger age at sexual debut (YASD) from the 31 studies that assessed YASD. (B) Forest plot illustrating pooled prevalence of YASD from the 31 studies grouped by sex of the study participants. (C) Forest plot illustrating pooled prevalence of YASD grouped by the African region in which the primary study was conducted. (D) Forest plot illustrating pooled prevalence of YASD grouped by year of publication of primary studies. (E) Forest plot illustrating pooled prevalence of YASD grouped by the definition of YASD in primary studies.

**S5 Fig. Forest plots showing pooled prevalence of concurrency**. (A) Forest plot illustrating pooled prevalence of concurrency from the 13 studies that assessed concurrency. (B) Forest plot illustrating pooled prevalence of concurrency from the 13 studies grouped by sex of the study participants. (C) Forest plot illustrating pooled prevalence of concurrency grouped by the African region in which the primary study was conducted. (D) Forest plot illustrating pooled prevalence of concurrency grouped by year of publication of primary studies. (E) Forest plot illustrating pooled prevalence of concurrency grouped by the definition of concurrency in primary studies.

**S6 Fig. Forest plots showing pooled prevalence of age-disparate relationships**. (A) Forest plot illustrating pooled prevalence of age-disparate relationships from the 12 studies that assessed age-disparate relationships. (B) Forest plot illustrating pooled prevalence of age-disparate relationships from the 12 studies grouped by sex of the study participants. (C) Forest plot illustrating pooled prevalence of age-disparate relationships grouped by the African region in which the primary study was conducted. (D) Forest plot illustrating pooled prevalence of age-disparate relationships grouped by year of publication of primary studies. (E) Forest plot illustrating pooled prevalence of age-disparate relationships grouped by the definition of age-disparate relationships in primary studies.

**S7 Fig. Forest plots showing pooled prevalence of study-defined SRTB**. (A) Forest plot illustrating pooled prevalence of study-defined sexual risk-taking behaviour (SRTB) from the 14 studies that assessed study-defined SRTB. (B) Forest plot illustrating pooled prevalence of study-defined SRTB from the 14 studies grouped by sex of the study participants. (C) Forest plot illustrating pooled prevalence of study-defined SRTB grouped by the African region in which the primary study was conducted. (D) Forest plot illustrating pooled prevalence of study-defined SRTB grouped by year of publication of primary studies. (E) Forest plot illustrating pooled prevalence of study-defined SRTB grouped by the definition of study-defined SRTB in primary studies.

## Acknowledgement

First, we would like to acknowledge the efforts of the scientists who authored the primary studies included in the current review. This work would not have been possible without their prior work. Secondly, we thank the researchers who availed their research articles upon request after we failed to obtain them in the various data bases. These include Dr. Mary B Adam (AIC Kijabe hospital, Kenya) and Dr. Yuichiro Yahata (Ministry of Health, Labour and Welfare, Japan). Finally, we appreciate Prof Hans Koot (Vrije Universiteit Amsterdam, Netherlands) and Mr. Alex Maina (Centre for Geographic Medicine Research, Kenya Medical Research Institute, Kenya) for their help during formal search for research articles.

