## Supplementary material for "Sexual risk-taking behavior amongst emerging adults in Africa: a systematic review and meta-analysis"

### S1 File: Full search syntax

(risk-taking behave* OR risky behav* OR risk behav*) AND (sexual* OR sexual risk OR unsafe sex OR sexual risk-taking OR reproductive OR sexual behav*) AND (emerging adult* OR college student* OR university student* OR young adult* OR youth* OR adolesce* OR young people) AND (Africa OR Africa* OR Algeria OR Angola OR Benin OR Botswana OR "Burkina Faso" OR Burundi OR Cameroon OR "Canary Islands" OR "Cape Verde" OR "Central African Republic" OR Chad OR Comoros OR Congo OR "Democratic Republic of Congo" OR Djibouti OR Egypt OR "Equatorial Guinea" OR Eritrea OR Ethiopia OR Gabon OR Gambia OR Ghana OR Guinea OR "Guinea Bissau" OR "Ivory Coast" OR "Cote d'Ivoire" OR Jamahiriya OR Jamahiryia OR Kenya OR Lesotho OR Liberia OR Libya OR Libia OR Madagascar OR Malawi OR Mali OR Mauritania OR Mauritius OR Mayote OR Morocco OR Mozambique OR Mocambique OR Namibia OR Niger OR Nigeria OR Principe OR Reunion OR Rwanda OR "Sao Tome" OR Senegal OR Seychelles OR "Sierra Leone" OR Somalia OR "South Africa" OR "St Helena" OR Sudan OR "South Sudan" OR Swaziland OR Tanzania OR Togo OR Tunisia OR Uganda OR "Western Sahara" OR Zaire OR Zambia OR Zimbabwe OR "Central Africa" OR "Central African" OR "West Africa" OR "West African" OR "Western Africa" OR "Western African" OR "East Africa" OR "East African" OR "Eastern Africa" OR "Eastern African" OR "North Africa" OR "North African" OR "Northern Africa" OR "Northern African" OR "South African" OR "Southern Africa" OR "Southern African" OR "sub Saharan Africa" OR "sub Saharan African" OR "sub-Saharan Africa" OR "sub-Saharan African") NOT ("guinea pig" OR "guinea pigs" OR "aspergillus niger").

### S2 File: PRISMA 2009 Checklist

| Section/topic | # | Checklist item | Reported on page # |
|---|---|---|---|
| TITLE | | | |
| Title | 1 | Identify the report as a systematic review, meta-analysis, or both. | 1 |
| ABSTRACT | | | |
| Structured summary | 2 | Provide a structured summary including, as applicable: background; objectives; data sources; study eligibility criteria, participants, and interventions; study appraisal and synthesis methods; results; limitations; conclusions and implications of key findings; systematic review registration number. | 2-3 |
| INTRODUCTION | | | |
| Rationale | 3 | Describe the rationale for the review in the context of what is already known. | 5 |
| Objectives | 4 | Provide an explicit statement of questions being addressed with reference to participants, interventions, comparisons, outcomes, and study design (PICOS). | 5 |
| METHODS | | | |
| Protocol and registration | 5 | Indicate if a review protocol exists, if and where it can be accessed (e.g., Web address), and, if available, provide registration information including registration number. | 6 |
| Eligibility criteria | 6 | Specify study characteristics (e.g., PICOS, length of follow-up) and report characteristics (e.g., years considered, language, publication status) used as criteria for eligibility, giving rationale. | 6 |
| Information sources | 7 | Describe all information sources (e.g., databases with dates of coverage, contact with study authors to identify additional studies) in the search and date last searched. | 6 |
| Search | 8 | Present full electronic search strategy for at least one database, including any limits used, such that it could be repeated. | S1_file.docx |
| Study selection | 9 | State the process for selecting studies (i.e., screening, eligibility, included in systematic review, and, if applicable, included in the meta-analysis). | 6 |
| Data collection process | 10 | Describe method of data extraction from reports (e.g., piloted forms, independently, in duplicate) and any processes for obtaining and confirming data from investigators. | 7 |
| Data items | 11 | List and define all variables for which data were sought (e.g., PICOS, funding sources) and any assumptions and simplifications made. | 6-7 |
| Risk of bias in individual studies | 12 | Describe methods used for assessing risk of bias of individual studies (including specification of whether this was done at the study or outcome level), and how this information is to be used in any data synthesis. | 7-8 |
| Summary measures | 13 | State the principal summary measures (e.g., risk ratio, difference in means). | 8-9 |
| Synthesis of results | 14 | Describe the methods of handling data and combining results of studies, if done, including measures of consistency (e.g., $I^2$) for each meta-analysis. | 8-9 |

| Section/topic | # | Checklist item | Reported on page # |
|---|---|---|---|
| Risk of bias across studies | 15 | Specify any assessment of risk of bias that may affect the cumulative evidence (e.g., publication bias, selective reporting within studies). | 7-8 |
| Additional analyses | 16 | Describe methods of additional analyses (e.g., sensitivity or subgroup analyses, meta-regression), if done, indicating which were pre-specified. | 8-9 |
| RESULTS | | | |
| Study selection | 17 | Give numbers of studies screened, assessed for eligibility, and included in the review, with reasons for exclusions at each stage, ideally with a flow diagram. | 10<br>Fig. 1 |
| Study characteristics | 18 | For each study, present characteristics for which data were extracted (e.g., study size, PICOS, follow-up period) and provide the citations. | 10<br>Table 1 |
| Risk of bias within studies | 19 | Present data on risk of bias of each study and, if available, any outcome level assessment (see item 12). | 10<br>Table 2 |
| Results of individual studies | 20 | For all outcomes considered (benefits or harms), present, for each study: (a) simple summary data for each intervention group (b) effect estimates and confidence intervals, ideally with a forest plot. | 12-19<br>S1-S7 Fig<br>Tables 3 and 4 |
| Synthesis of results | 21 | Present results of each meta-analysis done, including confidence intervals and measures of consistency. | 12-19<br>S1-S7 Fig<br>Tables 3 and 4 |
| Risk of bias across studies | 22 | Present results of any assessment of risk of bias across studies (see Item 15). | 10<br>Table 2 |
| Additional analysis | 23 | Give results of additional analyses, if done (e.g., sensitivity or subgroup analyses, meta-regression [see Item 16]). | N/A |
| DISCUSSION | | | |
| Summary of evidence | 24 | Summarize the main findings including the strength of evidence for each main outcome; consider their relevance to key groups (e.g., healthcare providers, users, and policy makers). | 19-23 |
| Limitations | 25 | Discuss limitations at study and outcome level (e.g., risk of bias), and at review-level (e.g., incomplete retrieval of identified research, reporting bias). | 23 |
| Conclusions | 26 | Provide a general interpretation of the results in the context of other evidence, and implications for future research. | 23-24 |
| FUNDING | | | |
| Funding | 27 | Describe sources of funding for the systematic review and other support (e.g., supply of data); role of funders for the systematic review. | In submission system |

**S1 Table: Definitions of SRTB outcomes as reported in primary studies**

| Outcome | Definition | No. of studies (%) | Total records (%) |
|---|---|---|---|
| Non- condom use | Condom non-use at last sex | 35 (37.6) | 42 (30.7) |
| | Condom non-use in last 12 months or less | 30 (32.3) | 33 (24.1) |
| | Condom non-use with no recall period | 23 (24.7) | 36 (26.3) |
| | Never used condom or non-use at first sex | 5 (5.4) | 26 (18.9) |
| | **Total** | **93** | **137** |
| Multiple sexual partnerships | MSP in last 12 months or less | 55 (79.7) | 57 (77.0) |
| | MSP with no recall period | 10 (14.5) | 10 (13.5) |
| | MSP in lifetime | 4 (5.8) | 7 (9.5) |
| | **Total** | **69** | **74** |
| Transactional sex | TS with no recall period | 17 (54.8) | 18 (56.3) |
| | TS in past 12 months or less | 9 (29.0) | 9 (28.1) |
| | TS in lifetime/ever | 5 (16.1) | 5 (15.6) |
| | **Total** | **31** | **32** |
| Younger age at sexual debut | First sex before 18 years of age | 26 (83.9) | 26 (83.9) |
| | First sex at age 18-20 years | 5 (16.1) | 5 (16.1) |
| | **Total** | **31** | **31** |
| Concurrency | Concurrency in last 12 months or less | 7 (53.9) | 7 (53.9) |
| | Current with unspecified recall period | 6 (46.1) | 6 (46.1) |
| | **Total** | **13** | **13** |
| Age-disparate relationships | Age difference 4 years or more | 10 (83.3) | 10 (83.3) |
| | Age difference unspecified | 2 (16.7) | 2 (16.7) |
| | **Total** | **12** | **12** |
| Study-defined SRTB | Self-defined SRTB in last 12 months or less | 8 (57.2) | 8 (57.2) |
| | Self-defined SRTB with no recall period | 3 (21.4) | 3 (21.4) |
| | Self-defined SRTB in lifetime/ever/last sex | 3 (21.4) | 3 (21.4) |
| | **Total** | **14** | **14** |

*MSP- Multiple sex partners; TS- Transactional sex; SRTB- Sexual-risk-taking behaviour

**S1 Figure:** (A) Forest plot illustrating pooled prevalence of non-condom use from the 137 records (obtained from 93 studies that assessed non-condom use). (B) Forest plot illustrating pooled prevalence of non-condom use grouped by sex of the study participants. (C) Forest plot illustrating pooled prevalence of non-condom use grouped by the African region in which the primary study was conducted. (D) Forest plot illustrating pooled prevalence of non-condom use grouped by the year of publication of primary studies. (E) Forest plot illustrating pooled prevalence of non-condom use grouped by the definition of non-condom use in primary studies.

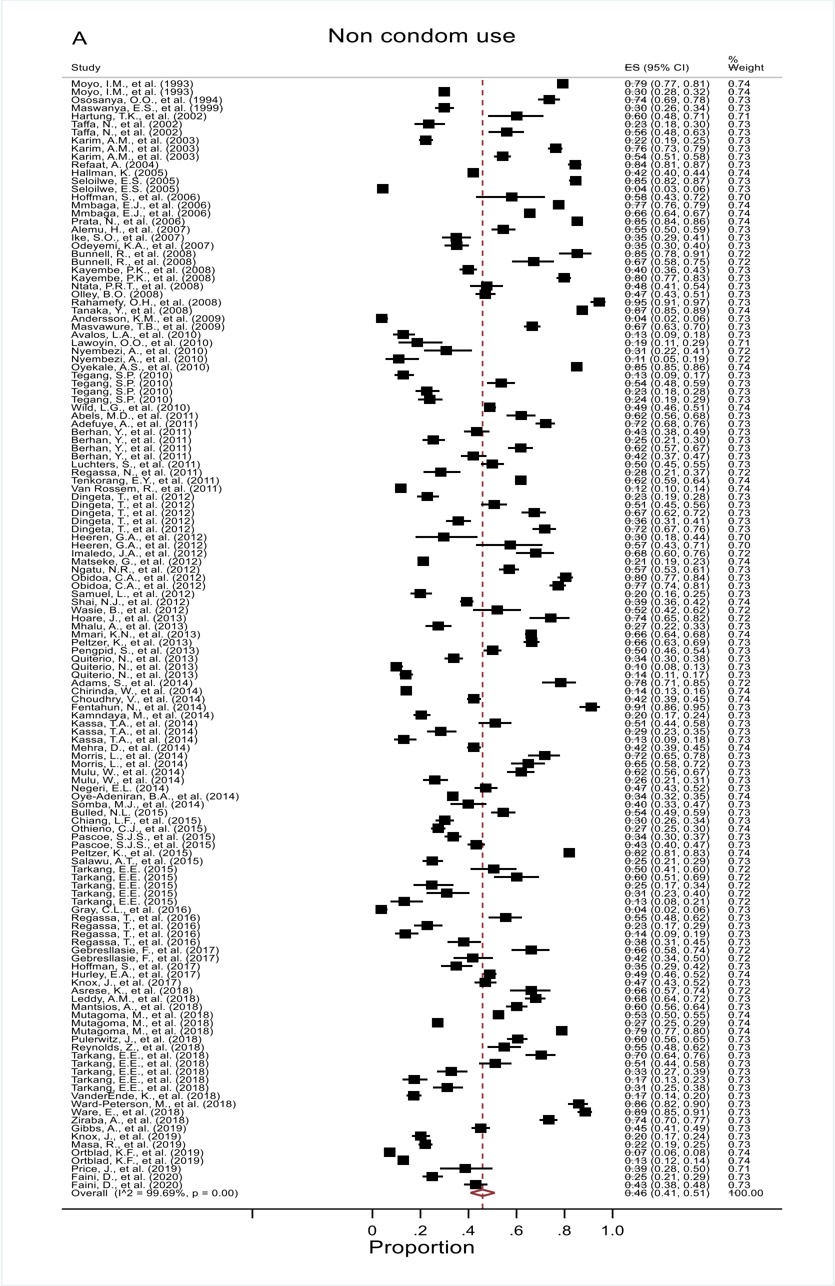

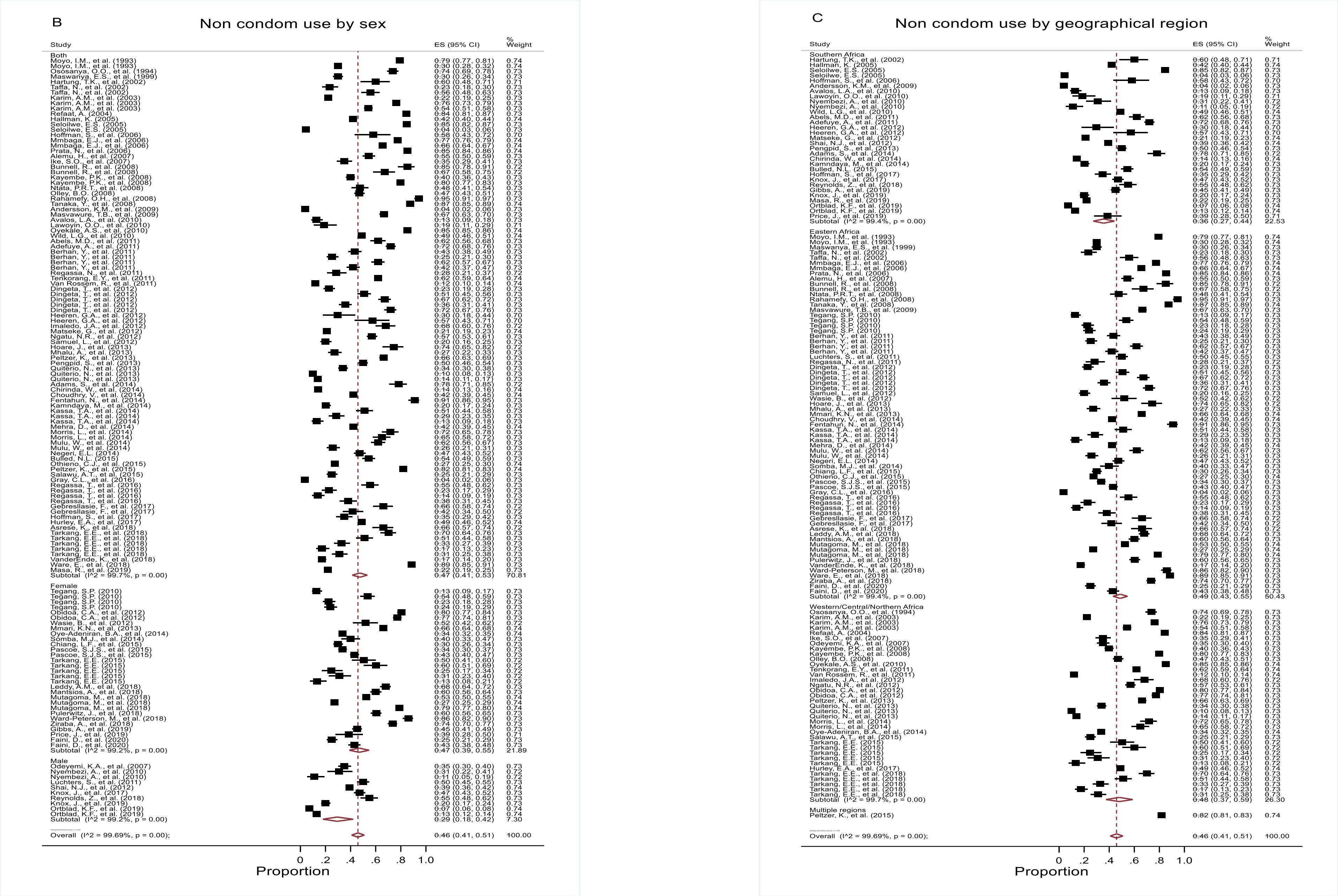
B
Non condom use by sex
Study
ES (95% CI)
% Weight
Both
Female
Male
Overall (I^2 = 99.69%, p = 0.00);
0 .2 .4 .6 .8 1.0
Proportion
C
Non condom use by geographical region
Study
ES (95% CI)
% Weight
Southern Africa
Eastern Africa
Western/Central/Northern Africa
Multiple regions
Overall (I^2 = 99.69%, p = 0.00);
0 .2 .4 .6 .8 1.0
Proportion

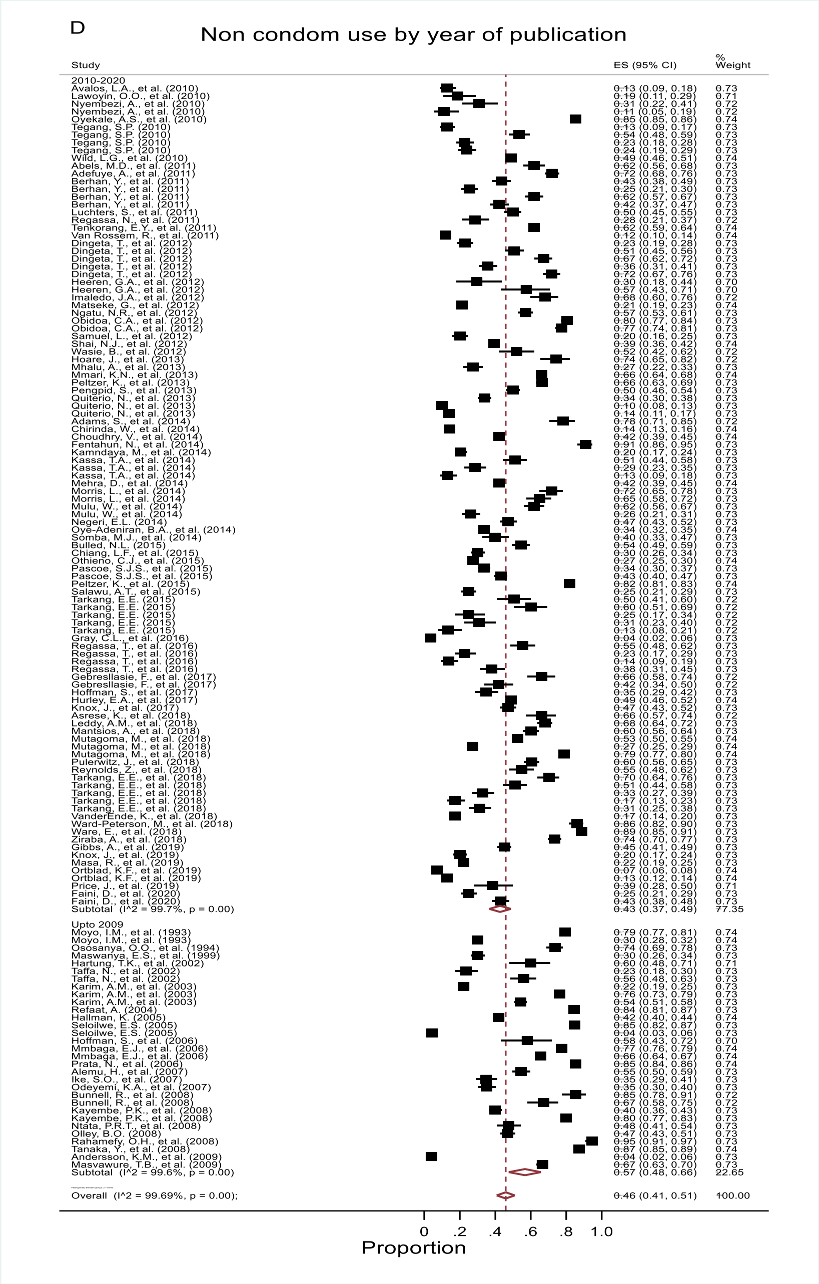
D
Non condom use by year of publication
Proportion

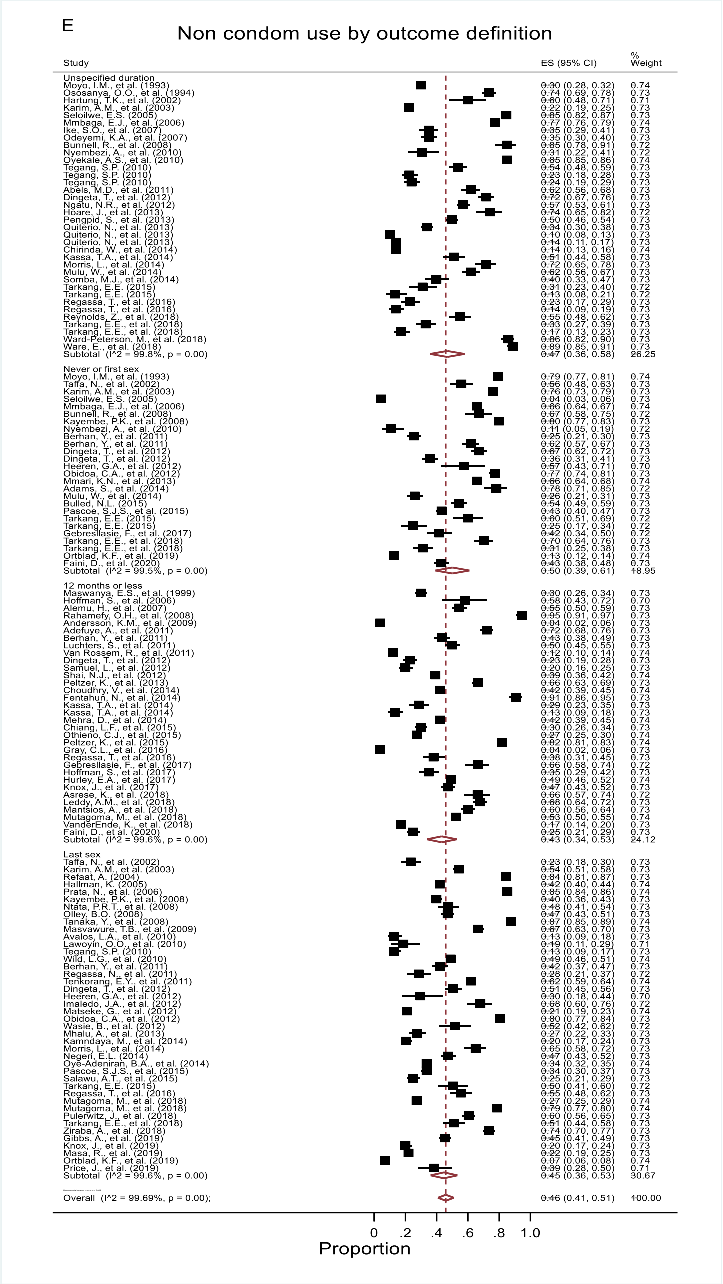
E
Non condom use by outcome definition
Proportion

**S2 Figure:** (A) Forest plot illustrating pooled prevalence of multiple sex partners (MSP) from the 74 records (obtained from 69 studies that assessed MSP). (B) Forest plot illustrating pooled prevalence of MSP from the 74 records grouped by sex of the study participants. (C) Forest plot illustrating pooled prevalence of MSP grouped by the African region in which the primary study was conducted. (D) Forest plot illustrating pooled prevalence of year of publication of primary studies. (E) Forest plot illustrating pooled prevalence of MSP grouped by the definition of MSP in primary studies.

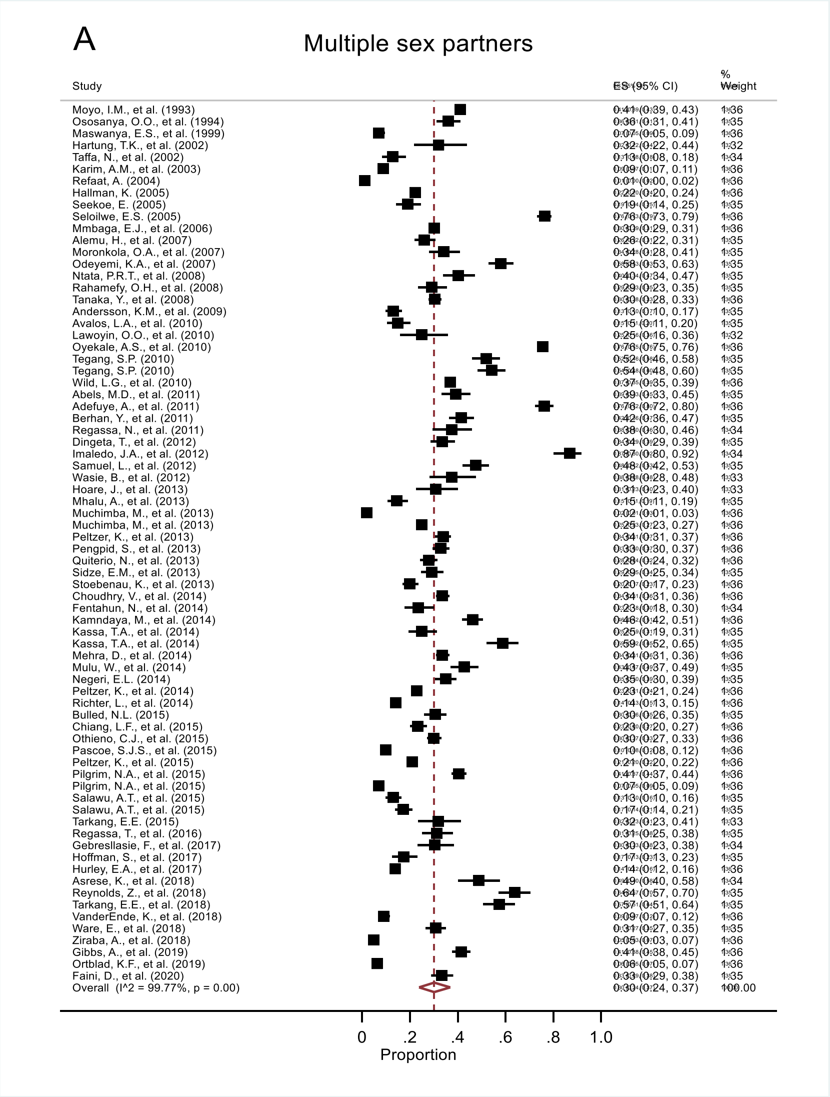

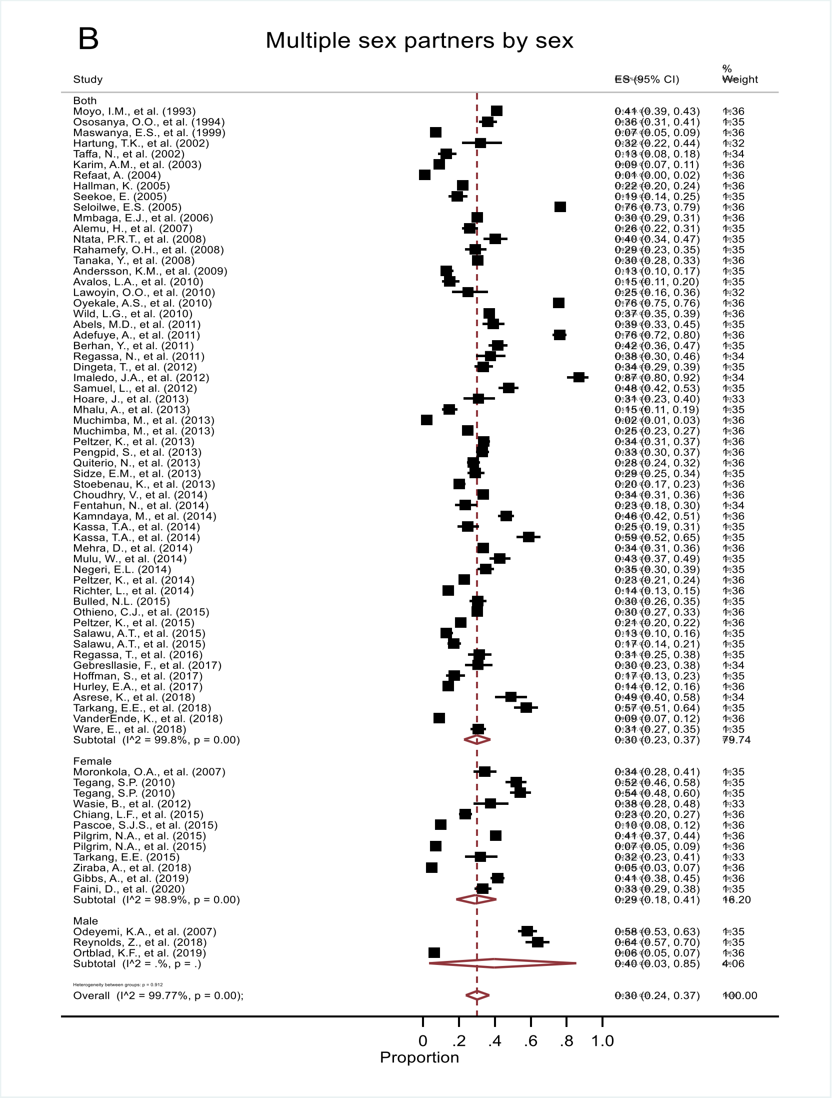
B
Multiple sex partners by sex
Study
ES (95% CI)
% Weight
Both
Moyo, I.M., et al. (1993) 0.41 (0.39, 0.43) 1.36
Ososanya, O.O., et al. (1994) 0.36 (0.31, 0.41) 1.35
Maswanya, E.S., et al. (1999) 0.07 (0.05, 0.09) 1.36
Hartung, T.K., et al. (2002) 0.32 (0.22, 0.44) 1.32
Taffa, N., et al. (2002) 0.13 (0.08, 0.18) 1.34
Karim, A.M., et al. (2003) 0.09 (0.07, 0.11) 1.36
Refaat, A. (2004) 0.01 (0.00, 0.02) 1.36
Hallman, K. (2005) 0.22 (0.20, 0.24) 1.36
Seekoe, E. (2005) 0.19 (0.14, 0.25) 1.35
Seloilwe, E.S. (2005) 0.76 (0.73, 0.79) 1.36
Mmbaga, E.J., et al. (2006) 0.30 (0.29, 0.31) 1.36
Alemu, H., et al. (2007) 0.26 (0.22, 0.31) 1.35
Ntata, P.R.T., et al. (2008) 0.40 (0.34, 0.47) 1.35
Rahamefy, O.H., et al. (2008) 0.29 (0.23, 0.35) 1.35
Tanaka, Y., et al. (2008) 0.30 (0.28, 0.33) 1.36
Andersson, K.M., et al. (2009) 0.13 (0.10, 0.17) 1.35
Avalos, L.A., et al. (2010) 0.15 (0.11, 0.20) 1.35
Lawoyin, O.O., et al. (2010) 0.25 (0.16, 0.36) 1.32
Oyekale, A.S., et al. (2010) 0.76 (0.75, 0.76) 1.36
Wild, L.G., et al. (2010) 0.37 (0.35, 0.39) 1.36
Abels, M.D., et al. (2011) 0.39 (0.33, 0.45) 1.35
Adefuye, A., et al. (2011) 0.76 (0.72, 0.80) 1.36
Berhan, Y., et al. (2011) 0.42 (0.36, 0.47) 1.35
Regassa, N., et al. (2011) 0.38 (0.30, 0.46) 1.34
Dingeta, T., et al. (2012) 0.34 (0.29, 0.39) 1.35
Imaledo, J.A., et al. (2012) 0.87 (0.80, 0.92) 1.34
Samuel, L., et al. (2012) 0.48 (0.42, 0.53) 1.35
Hoare, J., et al. (2013) 0.31 (0.23, 0.40) 1.33
Mhalu, A., et al. (2013) 0.15 (0.11, 0.19) 1.35
Muchimba, M., et al. (2013) 0.02 (0.01, 0.03) 1.36
Muchimba, M., et al. (2013) 0.25 (0.23, 0.27) 1.36
Peltzer, K., et al. (2013) 0.34 (0.31, 0.37) 1.36
Pengpid, S., et al. (2013) 0.33 (0.30, 0.37) 1.36
Quiterio, N., et al. (2013) 0.28 (0.24, 0.32) 1.36
Sidze, E.M., et al. (2013) 0.29 (0.25, 0.34) 1.35
Stoebenau, K., et al. (2013) 0.20 (0.17, 0.23) 1.36
Choudhry, V., et al. (2014) 0.34 (0.31, 0.36) 1.36
Fentahun, N., et al. (2014) 0.23 (0.18, 0.30) 1.34
Kamndaya, M., et al. (2014) 0.46 (0.42, 0.51) 1.36
Kassa, T.A., et al. (2014) 0.25 (0.19, 0.31) 1.35
Kassa, T.A., et al. (2014) 0.59 (0.52, 0.65) 1.35
Mehra, D., et al. (2014) 0.34 (0.31, 0.36) 1.36
Mulu, W., et al. (2014) 0.43 (0.37, 0.49) 1.35
Negeri, E.L. (2014) 0.35 (0.30, 0.39) 1.35
Peltzer, K., et al. (2014) 0.23 (0.21, 0.24) 1.36
Richter, L., et al. (2014) 0.14 (0.13, 0.15) 1.36
Bulled, N.L. (2015) 0.30 (0.26, 0.35) 1.35
Othieno, C.J., et al. (2015) 0.30 (0.27, 0.33) 1.36
Peltzer, K., et al. (2015) 0.21 (0.20, 0.22) 1.36
Salawu, A.T., et al. (2015) 0.13 (0.10, 0.16) 1.35
Salawu, A.T., et al. (2015) 0.17 (0.14, 0.21) 1.35
Regassa, T., et al. (2016) 0.31 (0.25, 0.38) 1.35
Gebreslasie, F., et al. (2017) 0.30 (0.23, 0.38) 1.34
Hoffman, S., et al. (2017) 0.17 (0.13, 0.23) 1.35
Hurley, E.A., et al. (2017) 0.14 (0.12, 0.16) 1.36
Asrese, K., et al. (2018) 0.49 (0.40, 0.58) 1.34
Tarkang, E.E., et al. (2018) 0.57 (0.51, 0.64) 1.35
VanderEnde, K., et al. (2018) 0.09 (0.07, 0.12) 1.36
Ware, E., et al. (2018) 0.31 (0.27, 0.35) 1.35
Subtotal (I^2 = 99.8%, p = 0.00) 0.30 (0.23, 0.37) 79.74
Female
Moronkola, O.A., et al. (2007) 0.34 (0.28, 0.41) 1.35
Tegang, S.P. (2010) 0.52 (0.46, 0.58) 1.35
Tegang, S.P. (2010) 0.54 (0.48, 0.60) 1.35
Wasie, B., et al. (2012) 0.38 (0.28, 0.48) 1.33
Chiang, L.F., et al. (2015) 0.23 (0.20, 0.27) 1.36
Pascoe, S.J.S., et al. (2015) 0.10 (0.08, 0.12) 1.36
Pilgrim, N.A., et al. (2015) 0.41 (0.37, 0.44) 1.36
Pilgrim, N.A., et al. (2015) 0.07 (0.05, 0.09) 1.36
Tarkang, E.E. (2015) 0.32 (0.23, 0.41) 1.33
Ziraba, A., et al. (2018) 0.05 (0.03, 0.07) 1.36
Gibbs, A., et al. (2019) 0.41 (0.38, 0.45) 1.36
Faini, D., et al. (2020) 0.33 (0.29, 0.38) 1.35
Subtotal (I^2 = 98.9%, p = 0.00) 0.29 (0.18, 0.41) 16.20
Male
Odeyemi, K.A., et al. (2007) 0.58 (0.53, 0.63) 1.35
Reynolds, Z., et al. (2018) 0.64 (0.57, 0.70) 1.35
Ortblad, K.F., et al. (2019) 0.06 (0.05, 0.07) 1.36
Subtotal (I^2 = .%, p = .) 0.40 (0.03, 0.85) 4.06
Overall (I^2 = 99.77%, p = 0.00); 0.30 (0.24, 0.37) 100.00
0 .2 .4 .6 .8 1.0
Proportion

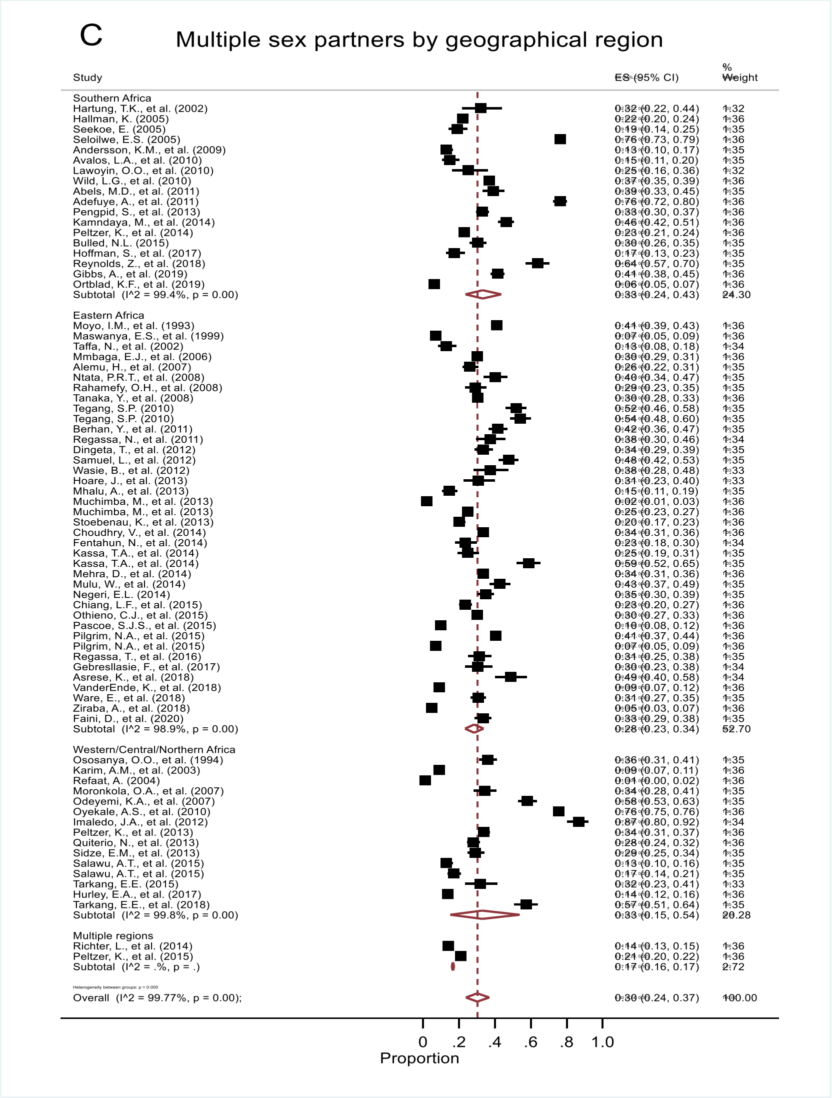
C
Multiple sex partners by geographical region
Study
ES (95% CI)
% Weight
Southern Africa
Hartung, T.K., et al. (2002) 0.32 (0.22, 0.44) 1.32
Hallman, K. (2005) 0.22 (0.20, 0.24) 1.36
Seekoe, E. (2005) 0.19 (0.14, 0.25) 1.35
Seloilwe, E.S. (2005) 0.76 (0.73, 0.79) 1.36
Andersson, K.M., et al. (2009) 0.13 (0.10, 0.17) 1.35
Avalos, L.A., et al. (2010) 0.15 (0.11, 0.20) 1.35
Lawoyin, O.O., et al. (2010) 0.25 (0.16, 0.36) 1.32
Wild, L.G., et al. (2010) 0.37 (0.35, 0.39) 1.36
Abels, M.D., et al. (2011) 0.39 (0.33, 0.45) 1.35
Adefuye, A., et al. (2011) 0.76 (0.72, 0.80) 1.36
Pengpid, S., et al. (2013) 0.33 (0.30, 0.37) 1.36
Kamndaya, M., et al. (2014) 0.46 (0.42, 0.51) 1.36
Peltzer, K., et al. (2014) 0.23 (0.21, 0.24) 1.36
Bulled, N.L. (2015) 0.30 (0.26, 0.35) 1.35
Hoffman, S., et al. (2017) 0.17 (0.13, 0.23) 1.35
Reynolds, Z., et al. (2018) 0.64 (0.57, 0.70) 1.35
Gibbs, A., et al. (2019) 0.41 (0.38, 0.45) 1.36
Ortblad, K.F., et al. (2019) 0.06 (0.05, 0.07) 1.36
Subtotal (I^2 = 99.4%, p = 0.00) 0.33 (0.24, 0.43) 24.30
Eastern Africa
Moyo, I.M., et al. (1993) 0.41 (0.39, 0.43) 1.36
Maswanya, E.S., et al. (1999) 0.07 (0.05, 0.09) 1.36
Taffa, N., et al. (2002) 0.13 (0.08, 0.18) 1.34
Mmbaga, E.J., et al. (2006) 0.30 (0.29, 0.31) 1.36
Alemu, H., et al. (2007) 0.26 (0.22, 0.31) 1.35
Ntata, P.R.T., et al. (2008) 0.40 (0.34, 0.47) 1.35
Rahamefy, O.H., et al. (2008) 0.29 (0.23, 0.35) 1.35
Tanaka, Y., et al. (2008) 0.30 (0.28, 0.33) 1.36
Tegang, S.P. (2010) 0.52 (0.46, 0.58) 1.35
Tegang, S.P. (2010) 0.54 (0.48, 0.60) 1.35
Berhan, Y., et al. (2011) 0.42 (0.36, 0.47) 1.35
Regassa, N., et al. (2011) 0.38 (0.30, 0.46) 1.34
Dingeta, T., et al. (2012) 0.34 (0.29, 0.39) 1.35
Samuel, L., et al. (2012) 0.48 (0.42, 0.53) 1.35
Wasie, B., et al. (2012) 0.38 (0.28, 0.48) 1.33
Hoare, J., et al. (2013) 0.31 (0.23, 0.40) 1.33
Mhalu, A., et al. (2013) 0.15 (0.11, 0.19) 1.35
Muchimba, M., et al. (2013) 0.02 (0.01, 0.03) 1.36
Muchimba, M., et al. (2013) 0.25 (0.23, 0.27) 1.36
Stoebenau, K., et al. (2013) 0.20 (0.17, 0.23) 1.36
Choudhry, V., et al. (2014) 0.34 (0.31, 0.36) 1.36
Fentahun, N., et al. (2014) 0.23 (0.18, 0.30) 1.34
Kassa, T.A., et al. (2014) 0.25 (0.19, 0.31) 1.35
Kassa, T.A., et al. (2014) 0.59 (0.52, 0.65) 1.35
Mehra, D., et al. (2014) 0.34 (0.31, 0.36) 1.36
Mulu, W., et al. (2014) 0.43 (0.37, 0.49) 1.35
Negeri, E.L. (2014) 0.35 (0.30, 0.39) 1.35
Chiang, L.F., et al. (2015) 0.23 (0.20, 0.27) 1.36
Othieno, C.J., et al. (2015) 0.30 (0.27, 0.33) 1.36
Pascoe, S.J.S., et al. (2015) 0.10 (0.08, 0.12) 1.36
Pilgrim, N.A., et al. (2015) 0.41 (0.37, 0.44) 1.36
Pilgrim, N.A., et al. (2015) 0.07 (0.05, 0.09) 1.36
Regassa, T., et al. (2016) 0.31 (0.25, 0.38) 1.35
Gebreslasie, F., et al. (2017) 0.30 (0.23, 0.38) 1.34
Asrese, K., et al. (2018) 0.49 (0.40, 0.58) 1.34
VanderEnde, K., et al. (2018) 0.09 (0.07, 0.12) 1.36
Ware, E., et al. (2018) 0.31 (0.27, 0.35) 1.35
Ziraba, A., et al. (2018) 0.05 (0.03, 0.07) 1.36
Faini, D., et al. (2020) 0.33 (0.29, 0.38) 1.35
Subtotal (I^2 = 98.9%, p = 0.00) 0.28 (0.23, 0.34) 52.70
Western/Central/Northern Africa
Ososanya, O.O., et al. (1994) 0.36 (0.31, 0.41) 1.35
Karim, A.M., et al. (2003) 0.09 (0.07, 0.11) 1.36
Refaat, A. (2004) 0.01 (0.00, 0.02) 1.36
Moronkola, O.A., et al. (2007) 0.34 (0.28, 0.41) 1.35
Odeyemi, K.A., et al. (2007) 0.58 (0.53, 0.63) 1.35
Oyekale, A.S., et al. (2010) 0.76 (0.75, 0.76) 1.36
Imaledo, J.A., et al. (2012) 0.87 (0.80, 0.92) 1.34
Peltzer, K., et al. (2013) 0.34 (0.31, 0.37) 1.36
Quiterio, N., et al. (2013) 0.28 (0.24, 0.32) 1.36
Sidze, E.M., et al. (2013) 0.29 (0.25, 0.34) 1.35
Salawu, A.T., et al. (2015) 0.13 (0.10, 0.16) 1.35
Salawu, A.T., et al. (2015) 0.17 (0.14, 0.21) 1.35
Tarkang, E.E. (2015) 0.32 (0.23, 0.41) 1.33
Hurley, E.A., et al. (2017) 0.14 (0.12, 0.16) 1.36
Tarkang, E.E., et al. (2018) 0.57 (0.51, 0.64) 1.35
Subtotal (I^2 = 99.8%, p = 0.00) 0.33 (0.15, 0.54) 20.28
Multiple regions
Richter, L., et al. (2014) 0.14 (0.13, 0.15) 1.36
Peltzer, K., et al. (2015) 0.21 (0.20, 0.22) 1.36
Subtotal (I^2 = .%, p = .) 0.17 (0.16, 0.17) 2.72
Overall (I^2 = 99.77%, p = 0.00); 0.30 (0.24, 0.37) 100.00
0 .2 .4 .6 .8 1.0
Proportion

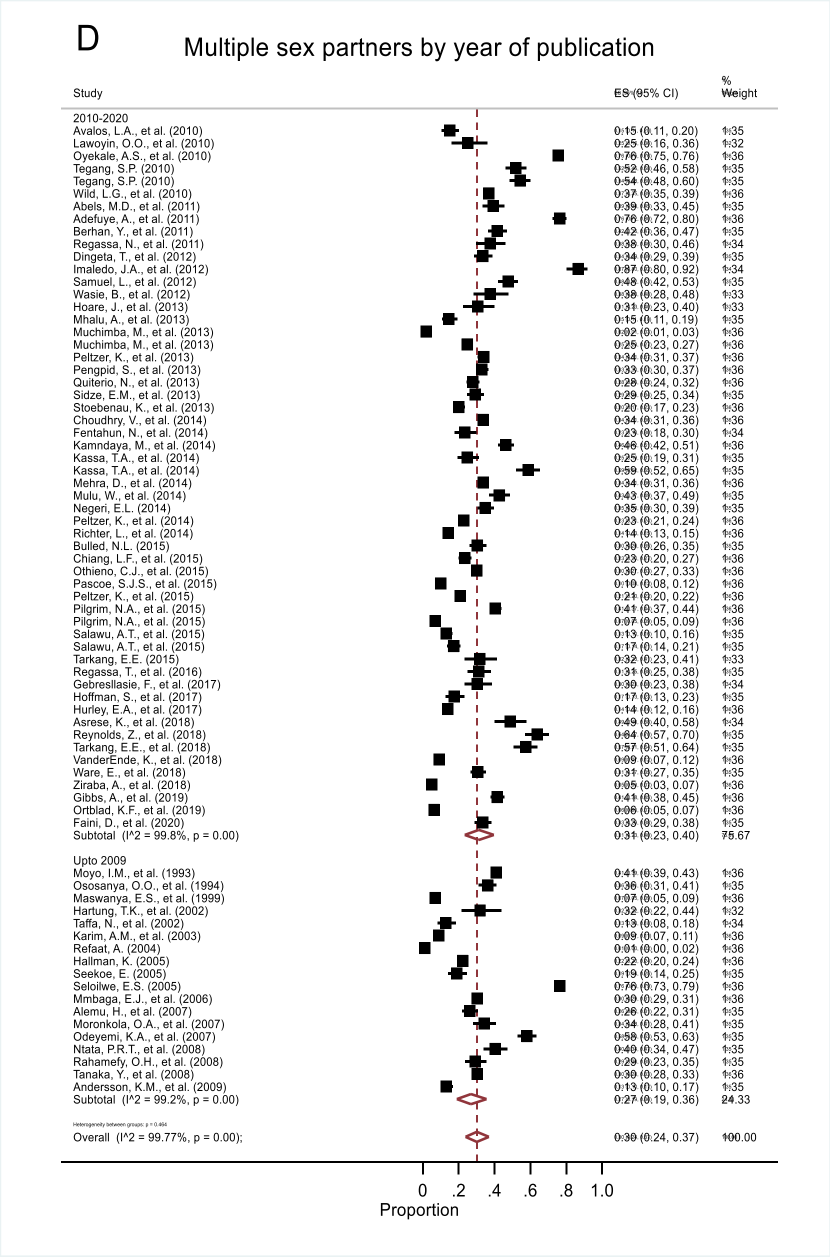
D
Multiple sex partners by year of publication
Study
ES (95% CI)
% Weight
2010-2020
Avalos, L.A., et al. (2010) 0.15 (0.11, 0.20) 1.35
Lawoyin, O.O., et al. (2010) 0.25 (0.16, 0.36) 1.32
Oyekale, A.S., et al. (2010) 0.76 (0.75, 0.76) 1.36
Tegang, S.P. (2010) 0.52 (0.46, 0.58) 1.35
Tegang, S.P. (2010) 0.54 (0.48, 0.60) 1.35
Wild, L.G., et al. (2010) 0.37 (0.35, 0.39) 1.36
Abels, M.D., et al. (2011) 0.39 (0.33, 0.45) 1.35
Adefuye, A., et al. (2011) 0.76 (0.72, 0.80) 1.36
Berhan, Y., et al. (2011) 0.42 (0.36, 0.47) 1.35
Regassa, N., et al. (2011) 0.38 (0.30, 0.46) 1.34
Dingeta, T., et al. (2012) 0.34 (0.29, 0.39) 1.35
Imaledo, J.A., et al. (2012) 0.87 (0.80, 0.92) 1.34
Samuel, L., et al. (2012) 0.48 (0.42, 0.53) 1.35
Wasie, B., et al. (2012) 0.38 (0.28, 0.48) 1.33
Hoare, J., et al. (2013) 0.31 (0.23, 0.40) 1.33
Mhalu, A., et al. (2013) 0.15 (0.11, 0.19) 1.35
Muchimba, M., et al. (2013) 0.02 (0.01, 0.03) 1.36
Muchimba, M., et al. (2013) 0.25 (0.23, 0.27) 1.36
Peltzer, K., et al. (2013) 0.34 (0.31, 0.37) 1.36
Pengpid, S., et al. (2013) 0.33 (0.30, 0.37) 1.36
Quiterio, N., et al. (2013) 0.28 (0.24, 0.32) 1.36
Sidze, E.M., et al. (2013) 0.29 (0.25, 0.34) 1.35
Stoebenau, K., et al. (2013) 0.20 (0.17, 0.23) 1.36
Choudhry, V., et al. (2014) 0.34 (0.31, 0.36) 1.36
Fentahun, N., et al. (2014) 0.23 (0.18, 0.30) 1.34
Kamndaya, M., et al. (2014) 0.46 (0.42, 0.51) 1.36
Kassa, T.A., et al. (2014) 0.25 (0.19, 0.31) 1.35
Kassa, T.A., et al. (2014) 0.59 (0.52, 0.65) 1.35
Mehra, D., et al. (2014) 0.34 (0.31, 0.36) 1.36
Mulu, W., et al. (2014) 0.43 (0.37, 0.49) 1.35
Negeri, E.L. (2014) 0.35 (0.30, 0.39) 1.35
Peltzer, K., et al. (2014) 0.23 (0.21, 0.24) 1.36
Richter, L., et al. (2014) 0.14 (0.13, 0.15) 1.36
Bulled, N.L. (2015) 0.30 (0.26, 0.35) 1.35
Chiang, L.F., et al. (2015) 0.23 (0.20, 0.27) 1.36
Othieno, C.J., et al. (2015) 0.30 (0.27, 0.33) 1.36
Pascoe, S.J.S., et al. (2015) 0.10 (0.08, 0.12) 1.36
Peltzer, K., et al. (2015) 0.21 (0.20, 0.22) 1.36
Pilgrim, N.A., et al. (2015) 0.41 (0.37, 0.44) 1.36
Pilgrim, N.A., et al. (2015) 0.07 (0.05, 0.09) 1.36
Salawu, A.T., et al. (2015) 0.13 (0.10, 0.16) 1.35
Salawu, A.T., et al. (2015) 0.17 (0.14, 0.21) 1.35
Tarkang, E.E. (2015) 0.32 (0.23, 0.41) 1.33
Regassa, T., et al. (2016) 0.31 (0.25, 0.38) 1.35
Gebresllasie, F., et al. (2017) 0.30 (0.23, 0.38) 1.34
Hoffman, S., et al. (2017) 0.17 (0.13, 0.23) 1.35
Hurley, E.A., et al. (2017) 0.14 (0.12, 0.16) 1.36
Asrese, K., et al. (2018) 0.49 (0.40, 0.58) 1.34
Reynolds, Z., et al. (2018) 0.64 (0.57, 0.70) 1.35
Tarkang, E.E., et al. (2018) 0.57 (0.51, 0.64) 1.35
VanderEnde, K., et al. (2018) 0.09 (0.07, 0.12) 1.36
Ware, E., et al. (2018) 0.31 (0.27, 0.35) 1.35
Ziraba, A., et al. (2018) 0.05 (0.03, 0.07) 1.36
Gibbs, A., et al. (2019) 0.41 (0.38, 0.45) 1.36
Ortblad, K.F., et al. (2019) 0.06 (0.05, 0.07) 1.36
Faini, D., et al. (2020) 0.33 (0.29, 0.38) 1.35
Subtotal (I^2 = 99.8%, p = 0.00) 0.31 (0.23, 0.40) 75.67
Upto 2009
Moyo, I.M., et al. (1993) 0.41 (0.39, 0.43) 1.36
Ososanya, O.O., et al. (1994) 0.36 (0.31, 0.41) 1.35
Maswanya, E.S., et al. (1999) 0.07 (0.05, 0.09) 1.36
Hartung, T.K., et al. (2002) 0.32 (0.22, 0.44) 1.32
Taffa, N., et al. (2002) 0.13 (0.08, 0.18) 1.34
Karim, A.M., et al. (2003) 0.09 (0.07, 0.11) 1.36
Refaat, A. (2004) 0.01 (0.00, 0.02) 1.36
Hallman, K. (2005) 0.22 (0.20, 0.24) 1.36
Seekoe, E. (2005) 0.19 (0.14, 0.25) 1.35
Seloilwe, E.S. (2005) 0.76 (0.73, 0.79) 1.36
Mmbaga, E.J., et al. (2006) 0.30 (0.29, 0.31) 1.36
Alemu, H., et al. (2007) 0.26 (0.22, 0.31) 1.35
Moronkola, O.A., et al. (2007) 0.34 (0.28, 0.41) 1.35
Odeyemi, K.A., et al. (2007) 0.58 (0.53, 0.63) 1.35
Ntata, P.R.T., et al. (2008) 0.40 (0.34, 0.47) 1.35
Rahamefy, O.H., et al. (2008) 0.29 (0.23, 0.35) 1.35
Tanaka, Y., et al. (2008) 0.30 (0.28, 0.33) 1.36
Andersson, K.M., et al. (2009) 0.13 (0.10, 0.17) 1.35
Subtotal (I^2 = 99.2%, p = 0.00) 0.27 (0.19, 0.36) 24.33
Overall (I^2 = 99.77%, p = 0.00); 0.30 (0.24, 0.37) 100.00
0 .2 .4 .6 .8 1.0
Proportion

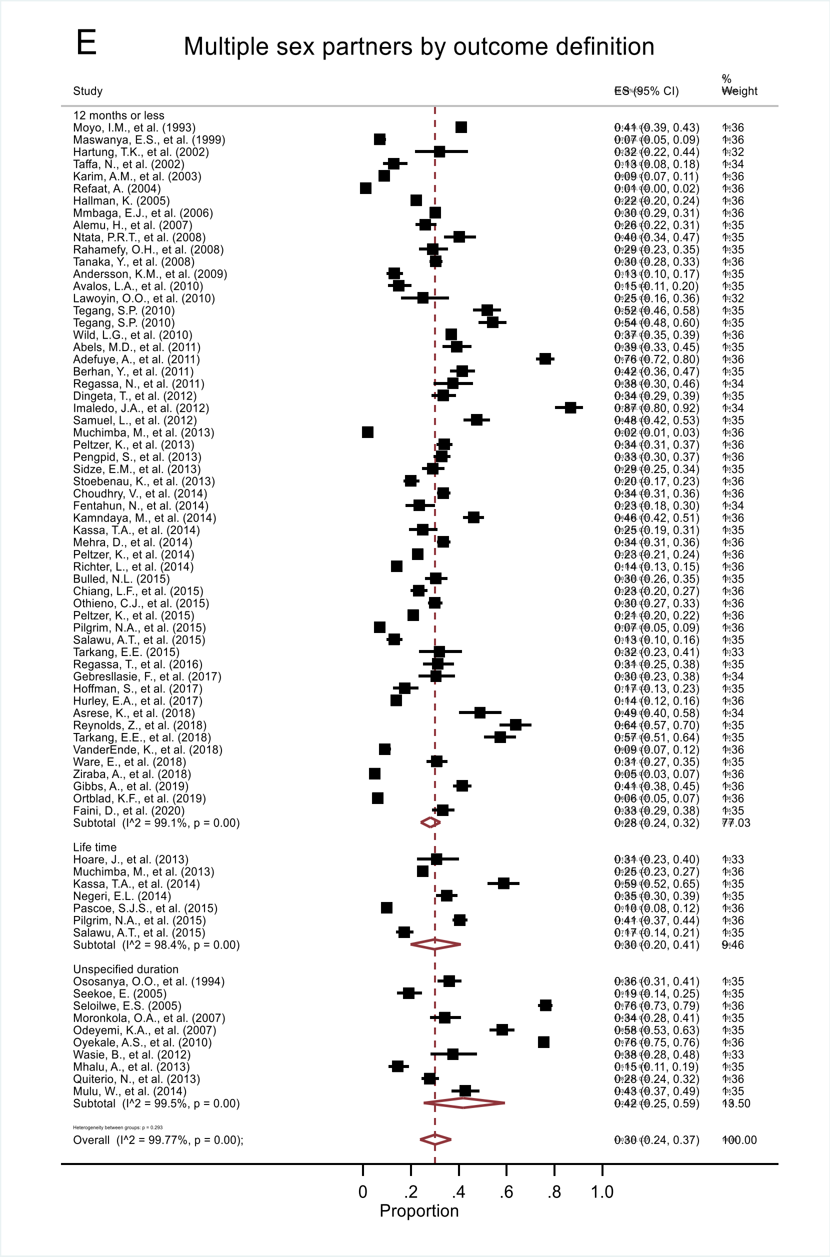
E
Multiple sex partners by outcome definition
Study
ES (95% CI)
% Weight
12 months or less
Moyo, I.M., et al. (1993) 0.41 (0.39, 0.43) 1.36
Maswanya, E.S., et al. (1999) 0.07 (0.05, 0.09) 1.36
Hartung, T.K., et al. (2002) 0.32 (0.22, 0.44) 1.32
Taffa, N., et al. (2002) 0.13 (0.08, 0.18) 1.34
Karim, A.M., et al. (2003) 0.09 (0.07, 0.11) 1.36
Refaat, A. (2004) 0.01 (0.00, 0.02) 1.36
Hallman, K. (2005) 0.22 (0.20, 0.24) 1.36
Mmbaga, E.J., et al. (2006) 0.30 (0.29, 0.31) 1.36
Alemu, H., et al. (2007) 0.26 (0.22, 0.31) 1.35
Ntata, P.R.T., et al. (2008) 0.40 (0.34, 0.47) 1.35
Rahamefy, O.H., et al. (2008) 0.29 (0.23, 0.35) 1.35
Tanaka, Y., et al. (2008) 0.30 (0.28, 0.33) 1.36
Andersson, K.M., et al. (2009) 0.13 (0.10, 0.17) 1.35
Avalos, L.A., et al. (2010) 0.15 (0.11, 0.20) 1.35
Lawoyin, O.O., et al. (2010) 0.25 (0.16, 0.36) 1.32
Tegang, S.P. (2010) 0.52 (0.46, 0.58) 1.35
Tegang, S.P. (2010) 0.54 (0.48, 0.60) 1.35
Wild, L.G., et al. (2010) 0.37 (0.35, 0.39) 1.36
Abels, M.D., et al. (2011) 0.39 (0.33, 0.45) 1.35
Adefuye, A., et al. (2011) 0.76 (0.72, 0.80) 1.36
Berhan, Y., et al. (2011) 0.42 (0.36, 0.47) 1.35
Regassa, N., et al. (2011) 0.38 (0.30, 0.46) 1.34
Dingeta, T., et al. (2012) 0.34 (0.29, 0.39) 1.35
Imaledo, J.A., et al. (2012) 0.87 (0.80, 0.92) 1.34
Samuel, L., et al. (2012) 0.48 (0.42, 0.53) 1.35
Muchimba, M., et al. (2013) 0.02 (0.01, 0.03) 1.36
Peltzer, K., et al. (2013) 0.34 (0.31, 0.37) 1.36
Pengpid, S., et al. (2013) 0.33 (0.30, 0.37) 1.36
Sidze, E.M., et al. (2013) 0.29 (0.25, 0.34) 1.35
Stoebenau, K., et al. (2013) 0.20 (0.17, 0.23) 1.36
Choudhry, V., et al. (2014) 0.34 (0.31, 0.36) 1.36
Fentahun, N., et al. (2014) 0.23 (0.18, 0.30) 1.34
Kamndaya, M., et al. (2014) 0.46 (0.42, 0.51) 1.36
Kassa, T.A., et al. (2014) 0.25 (0.19, 0.31) 1.35
Mehra, D., et al. (2014) 0.34 (0.31, 0.36) 1.36
Peltzer, K., et al. (2014) 0.23 (0.21, 0.24) 1.36
Richter, L., et al. (2014) 0.14 (0.13, 0.15) 1.36
Bulled, N.L. (2015) 0.30 (0.26, 0.35) 1.35
Chiang, L.F., et al. (2015) 0.23 (0.20, 0.27) 1.36
Othieno, C.J., et al. (2015) 0.30 (0.27, 0.33) 1.36
Peltzer, K., et al. (2015) 0.21 (0.20, 0.22) 1.36
Pilgrim, N.A., et al. (2015) 0.07 (0.05, 0.09) 1.36
Salawu, A.T., et al. (2015) 0.13 (0.10, 0.16) 1.35
Tarkang, E.E. (2015) 0.32 (0.23, 0.41) 1.33
Regassa, T., et al. (2016) 0.31 (0.25, 0.38) 1.35
Gebresllasie, F., et al. (2017) 0.30 (0.23, 0.38) 1.34
Hoffman, S., et al. (2017) 0.17 (0.13, 0.23) 1.35
Hurley, E.A., et al. (2017) 0.14 (0.12, 0.16) 1.36
Asrese, K., et al. (2018) 0.49 (0.40, 0.58) 1.34
Reynolds, Z., et al. (2018) 0.64 (0.57, 0.70) 1.35
Tarkang, E.E., et al. (2018) 0.57 (0.51, 0.64) 1.35
VanderEnde, K., et al. (2018) 0.09 (0.07, 0.12) 1.36
Ware, E., et al. (2018) 0.31 (0.27, 0.35) 1.35
Ziraba, A., et al. (2018) 0.05 (0.03, 0.07) 1.36
Gibbs, A., et al. (2019) 0.41 (0.38, 0.45) 1.36
Ortblad, K.F., et al. (2019) 0.06 (0.05, 0.07) 1.36
Faini, D., et al. (2020) 0.33 (0.29, 0.38) 1.35
Subtotal (I^2 = 99.1%, p = 0.00) 0.28 (0.24, 0.32) 77.03
Life time
Hoare, J., et al. (2013) 0.31 (0.23, 0.40) 1.33
Muchimba, M., et al. (2013) 0.25 (0.23, 0.27) 1.36
Kassa, T.A., et al. (2014) 0.59 (0.52, 0.65) 1.35
Negeri, E.L. (2014) 0.35 (0.30, 0.39) 1.35
Pascoe, S.J.S., et al. (2015) 0.10 (0.08, 0.12) 1.36
Pilgrim, N.A., et al. (2015) 0.41 (0.37, 0.44) 1.36
Salawu, A.T., et al. (2015) 0.17 (0.14, 0.21) 1.35
Subtotal (I^2 = 98.4%, p = 0.00) 0.30 (0.20, 0.41) 9.46
Unspecified duration
Ososanya, O.O., et al. (1994) 0.36 (0.31, 0.41) 1.35
Seekoe, E. (2005) 0.19 (0.14, 0.25) 1.35
Seloilwe, E.S. (2005) 0.76 (0.73, 0.79) 1.36
Moronkola, O.A., et al. (2007) 0.34 (0.28, 0.41) 1.35
Odeyemi, K.A., et al. (2007) 0.58 (0.53, 0.63) 1.35
Oyekale, A.S., et al. (2010) 0.76 (0.75, 0.76) 1.36
Wasie, B., et al. (2012) 0.38 (0.28, 0.48) 1.33
Mhalu, A., et al. (2013) 0.15 (0.11, 0.19) 1.35
Quiterio, N., et al. (2013) 0.28 (0.24, 0.32) 1.36
Mulu, W., et al. (2014) 0.43 (0.37, 0.49) 1.35
Subtotal (I^2 = 99.5%, p = 0.00) 0.42 (0.25, 0.59) 13.50
Overall (I^2 = 99.77%, p = 0.00); 0.30 (0.24, 0.37) 100.00
0 .2 .4 .6 .8 1.0
Proportion

**S3 Figure:** (A) Forest plot illustrating pooled prevalence of transactional sex (TS) from the 32 records (obtained from 31 studies that assessed TS). (B) Forest plot illustrating pooled prevalence of TS from the 32 records grouped by sex of the study participants. (C) Forest plot illustrating pooled prevalence of TS grouped by the African region in which the primary study was conducted. (D) Forest plot illustrating pooled prevalence of TS by year of publication of primary studies. (E) Forest plot illustrating pooled prevalence of TS grouped by the definition of TS in primary studies.

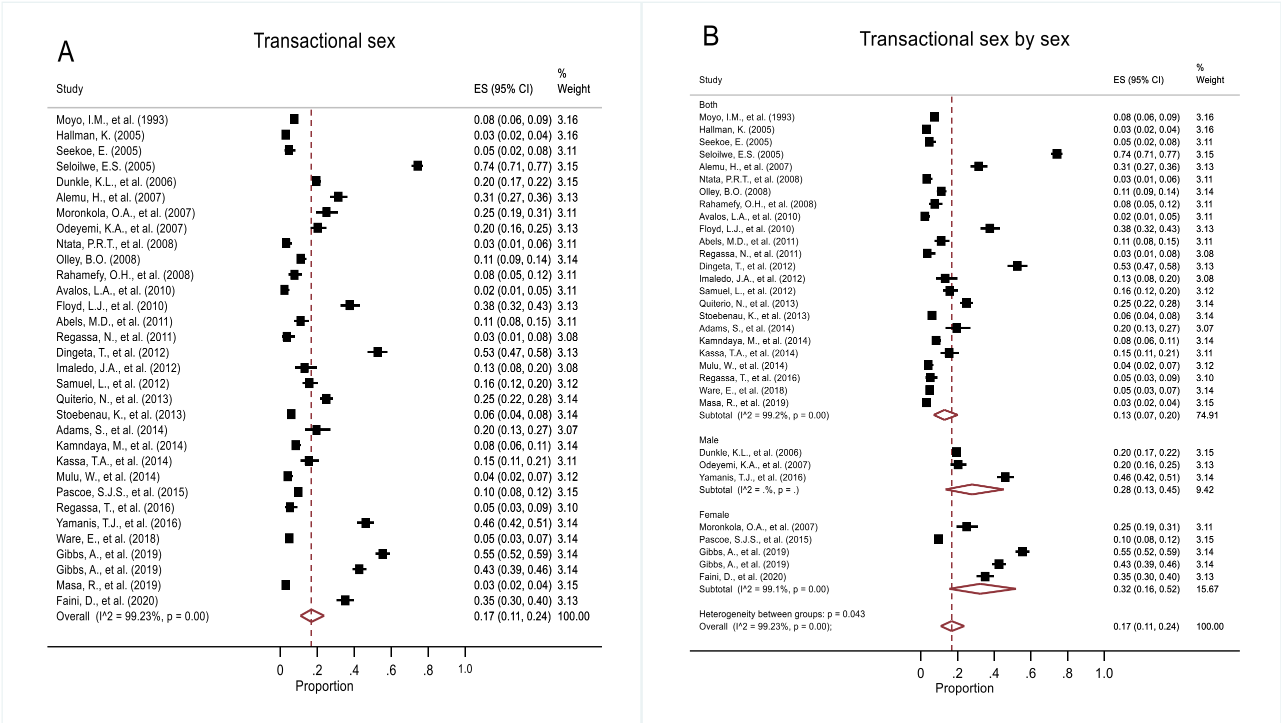


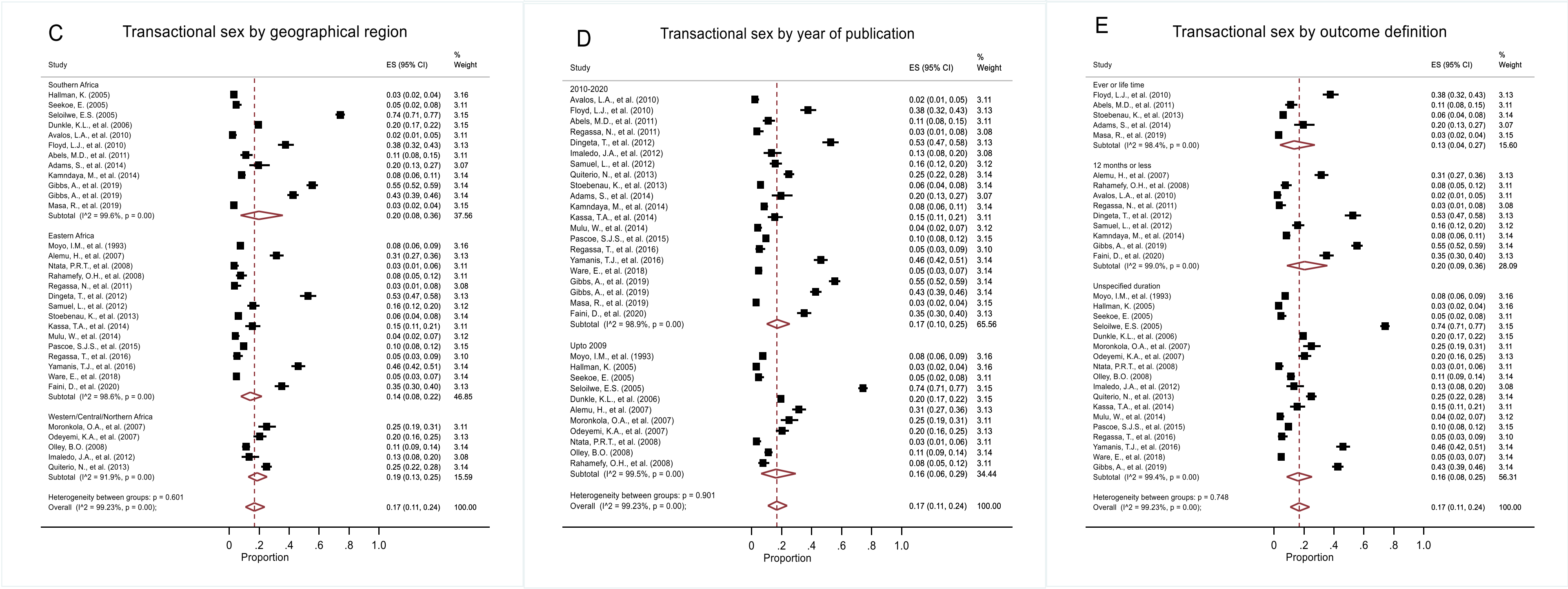

**S4 Figure:** (A) Forest plot illustrating pooled prevalence of younger age at sexual debut (YASD) from the 31 studies that assessed YASD. (B) Forest plot illustrating pooled prevalence of YASD from the 31 studies grouped by sex of the study participants. (C) Forest plot illustrating pooled prevalence of YASD grouped by the African region in which the primary study was conducted. (D) Forest plot illustrating pooled prevalence of YASD grouped by year of publication of primary studies. (E) Forest plot illustrating pooled prevalence of YASD grouped by the definition of YASD in primary studies.

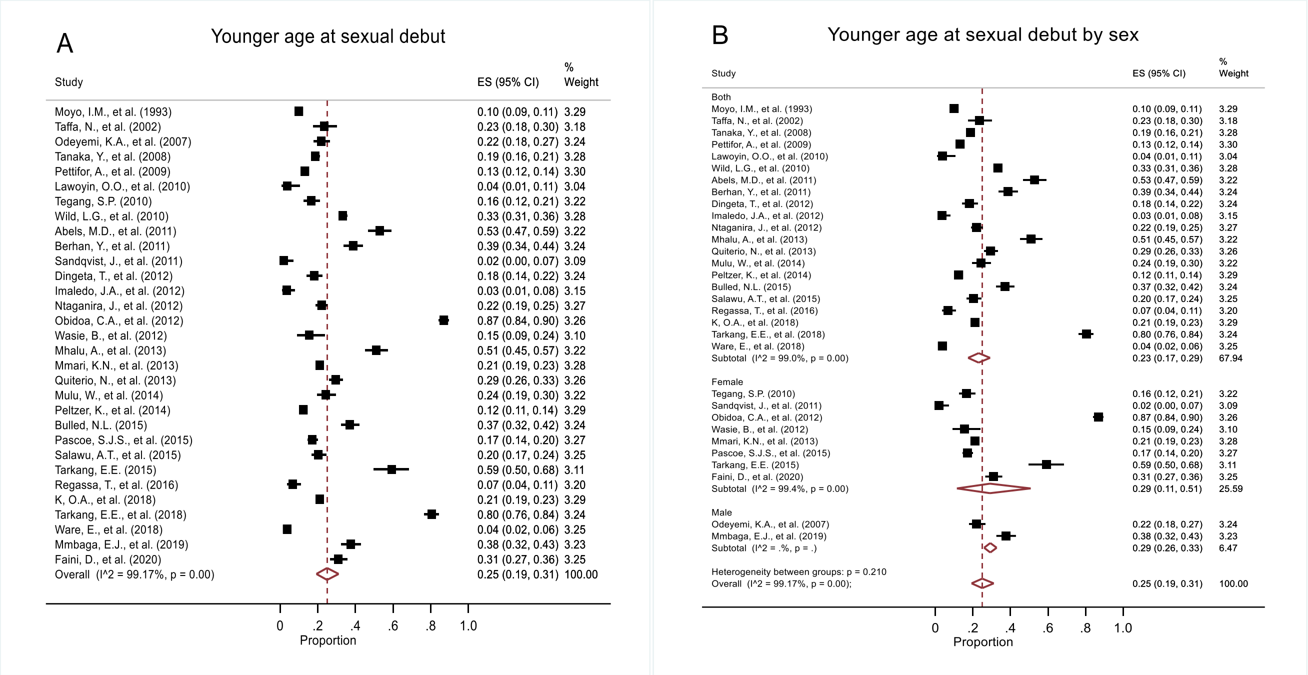


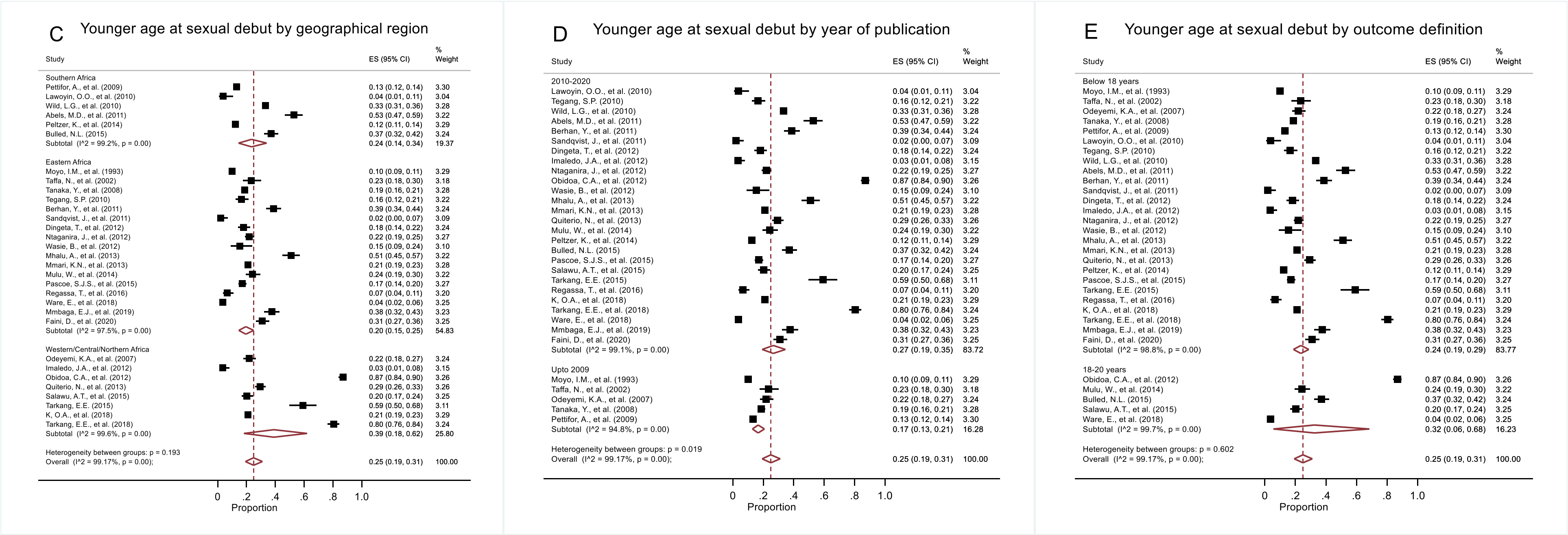

**S5 Figure:** (A) Forest plot illustrating pooled prevalence of concurrency from the 13 studies that assessed concurrency. (B) Forest plot illustrating pooled prevalence of concurrency from the 13 studies grouped by sex of the study participants. (C) Forest plot illustrating pooled prevalence of concurrency grouped by the African region in which the primary study was conducted. (D) Forest plot illustrating pooled prevalence of concurrency grouped by year of publication of primary studies. (E) Forest plot illustrating pooled prevalence of concurrency grouped by the definition of concurrency in primary studies.

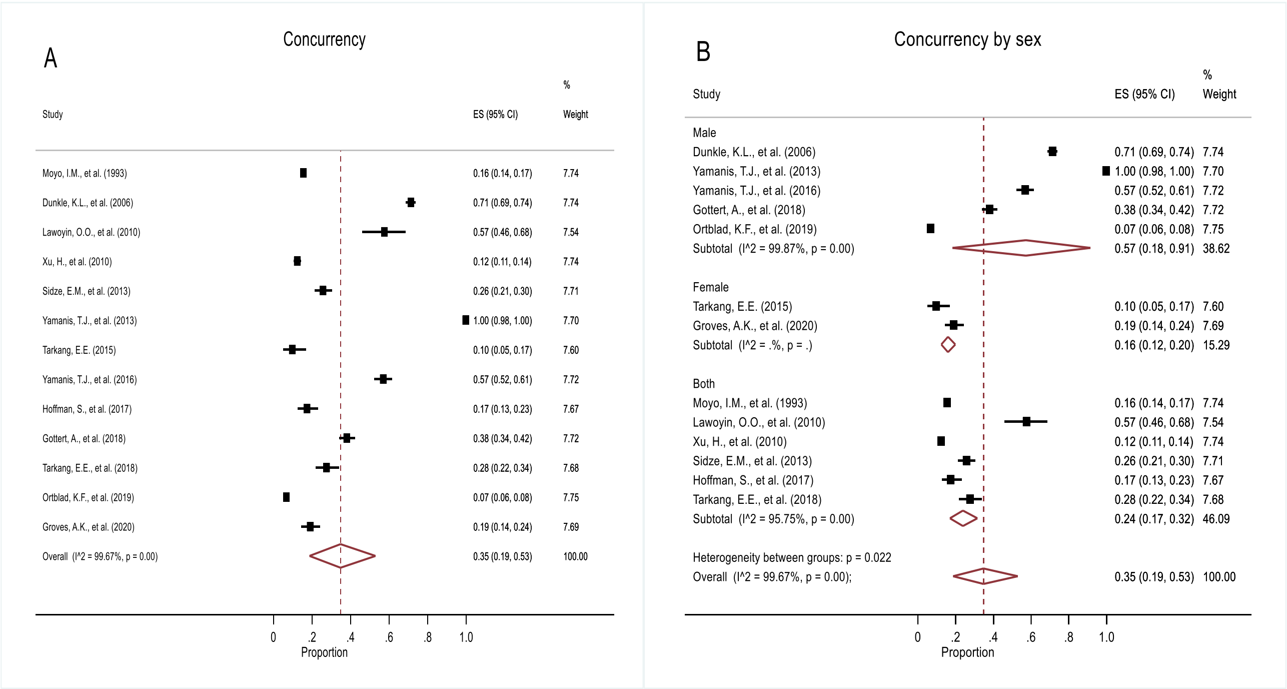


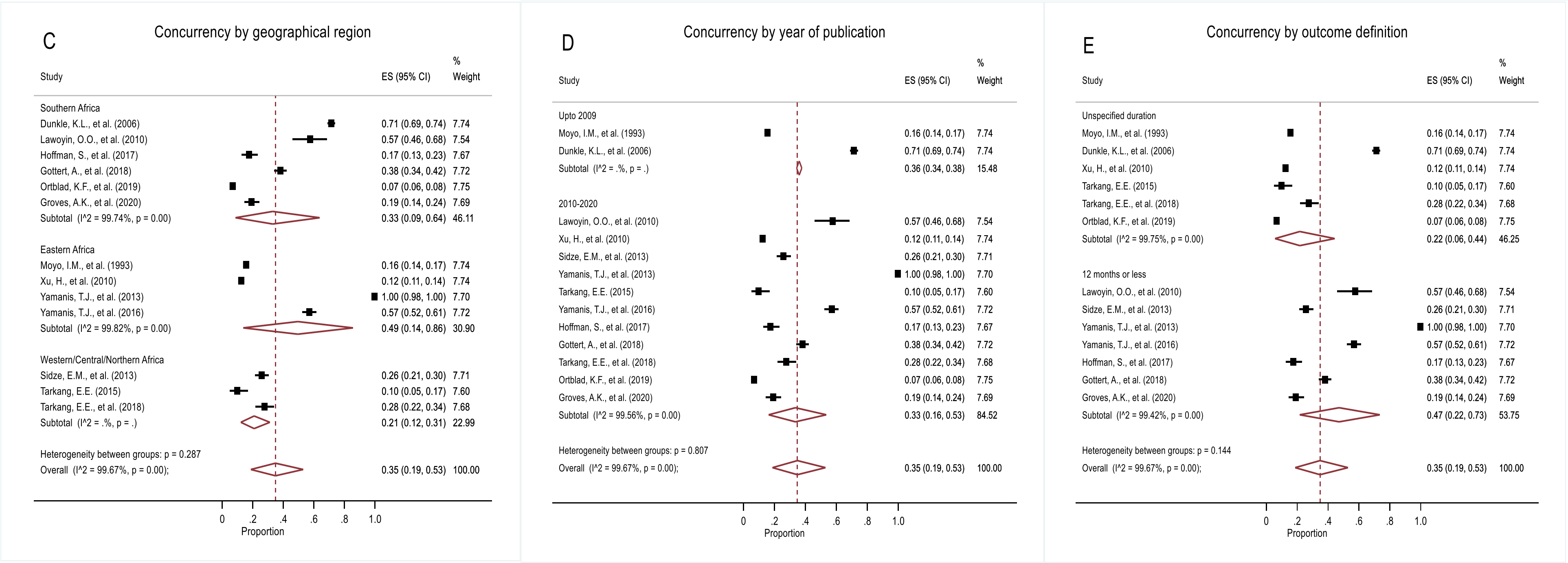

**S6 Figure:** (A) Forest plot illustrating pooled prevalence of age-disparate relationships from the 12 studies that assessed age-disparate relationships. (B) Forest plot illustrating pooled prevalence of age-disparate relationships from the 12 studies grouped by sex of the study participants. (C) Forest plot illustrating pooled prevalence of age-disparate relationships grouped by the African region in which the primary study was conducted. (D) Forest plot illustrating pooled prevalence of age-disparate relationships grouped by year of publication of primary studies. (E) Forest plot illustrating pooled prevalence of age-disparate relationships grouped by the definition of age-disparate relationships in primary studies.

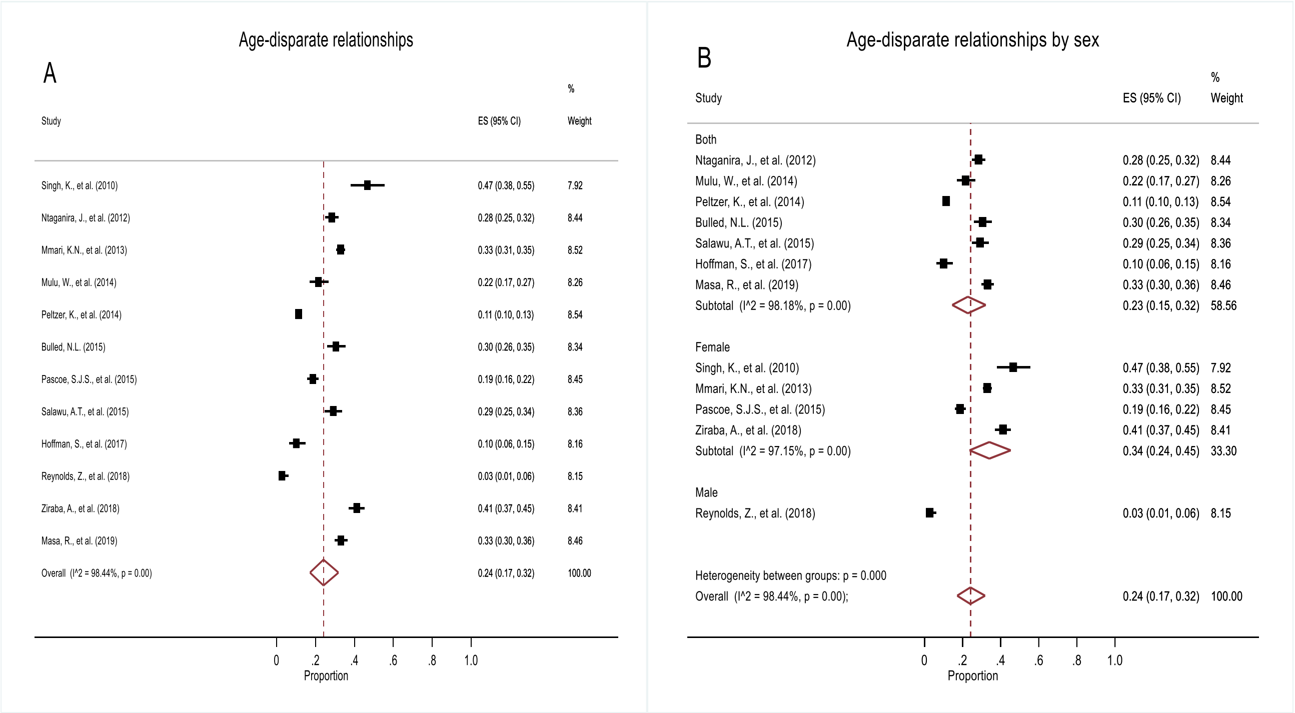


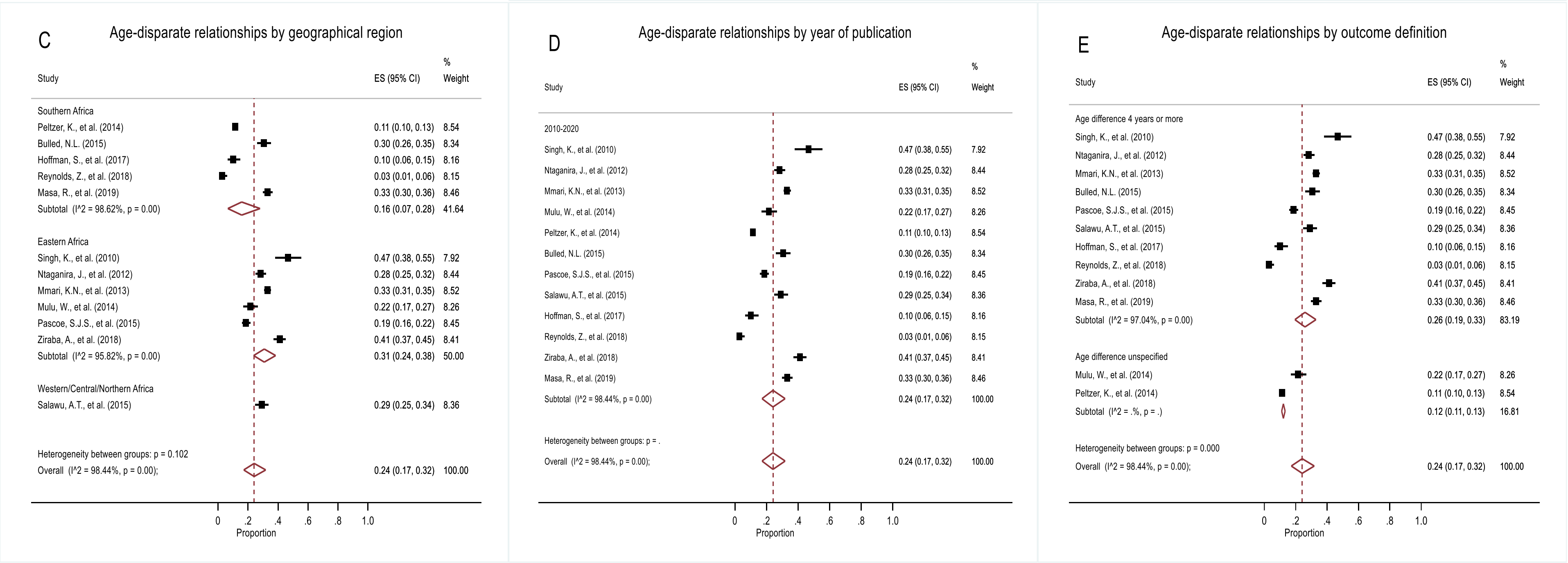

**S7 Figure:** (A) Forest plot illustrating pooled prevalence of study-defined sexual risk-taking behaviour (SRTB) from the 14 studies that assessed study-defined SRTB. (B) Forest plot illustrating pooled prevalence of study-defined SRTB from the 14 studies grouped by sex of the study participants. (C) Forest plot illustrating pooled prevalence of study-defined SRTB grouped by the African region in which the primary study was conducted. (D) Forest plot illustrating pooled prevalence of study-defined SRTB grouped by year of publication of primary studies. (E) Forest plot illustrating pooled prevalence of study-defined SRTB grouped by the definition of study-defined SRTB in primary studies.

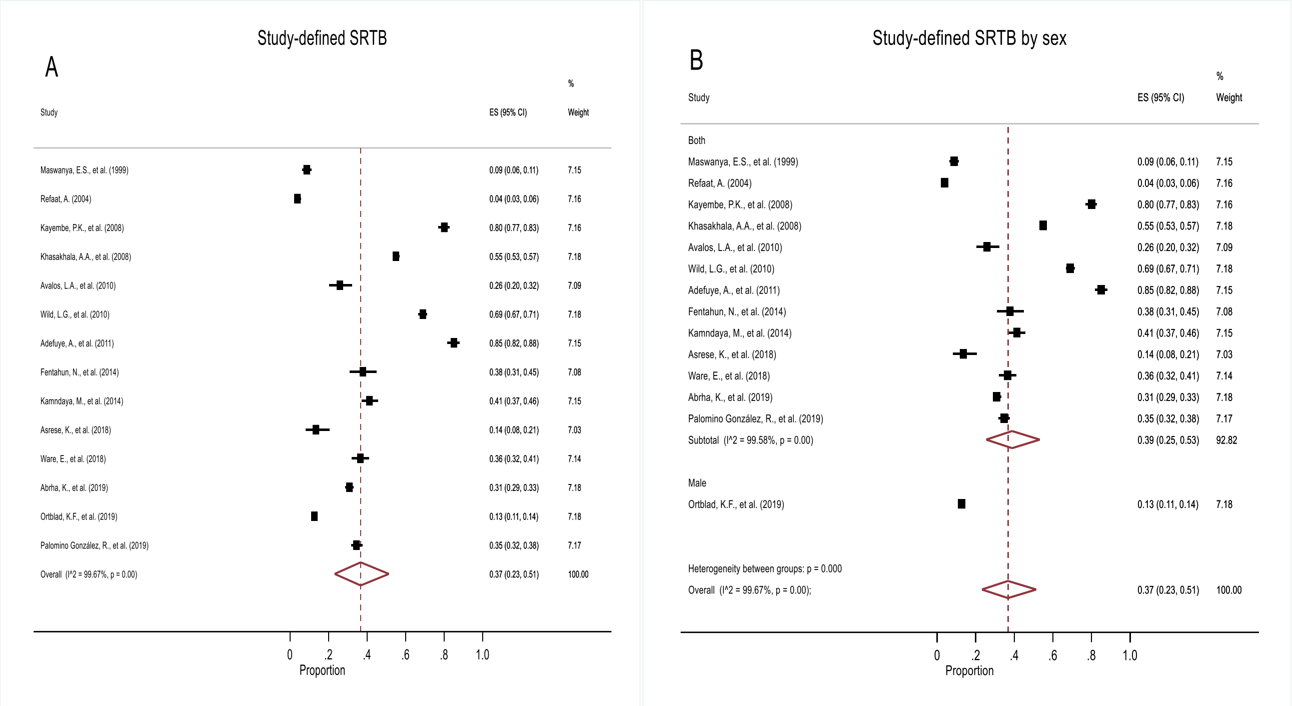


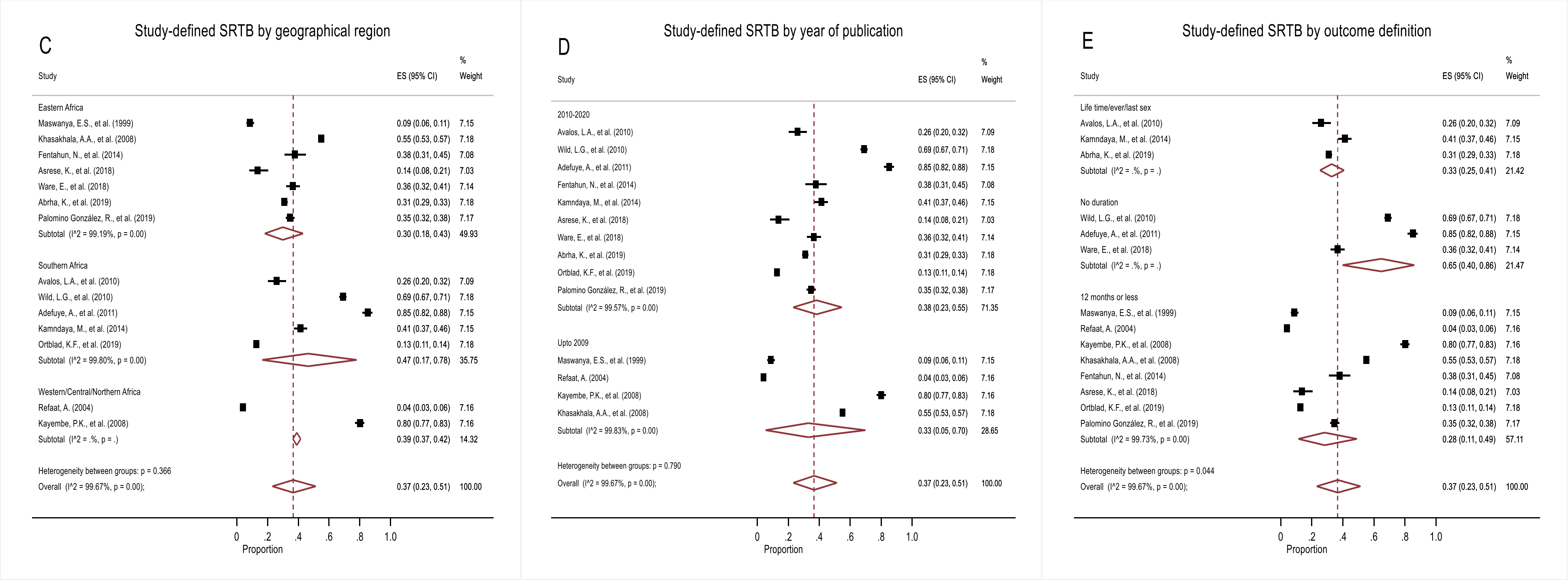

**S2 Table: Summary of factors assessed for their association with individual SRTB outcomes in studies included in analysis grouped in five thematic areas derived from a modified ecological model**

| | Non- condom use | Multiple sex partners | Transactional sex | Younger age at sexual debut | Concurrency | Age-disparate relationships | Study-defined SRTB |
|---|---|---|---|---|---|---|---|
| Socio-demographic factors | Age (n=47); education (n=38); socioeconomic status (n=26); residence (n=24); marital status (n=21); gender (n=20); religion (n=18); ethnicity (n=5); number of children (n=1) | Age (n=28); education (n=25); gender (n=16); residence (n=14); marital status (n=13); socioeconomic status (n=13; religion (n=13); ethnicity (n=5) | Age (n=11); education (n=9); socioeconomic status (n=6); marital status (n=4); ethnicity (n=3); residence (n=3); religion (n=2) | Age (n=9); education (n=9); socioeconomic status (n=8); religion (n=5); marital status (n=3); residence (n=3); ethnicity (n=2); gender (n=2) | Age (n=9); education (n=7); socioeconomic status (n=5); marital status (n=4); religion (n=2); gender (n=1); residence (n=1); where slept last night (n=1) | Age (n=3); socioeconomic status/food security (n=3); education (n=2); marital status (n=2); gender (n=1); age 1st married (n=1); ethnicity (n=1); residence (n=1); religion (n=1) | Age (n=11); education (n=10); sex (n=7); socioeconomic status (n=7); residence (n=9); marital status (n=2); religion (n=4); ethnicity (n=2) |
| Relationship/behavioural factors | Drug/alcohol use(n=30); sexual/physical violence/abuse (n=18); number of sex partners (n=11); sex debut (n=10); transactional sex (n=8); partner type/characteristics (n=8); having sex partner/sexually active (n=7); relationship power/control (n=6); peer pressure (n=6); talking to partner about condom/RH issues (n=5); watching porn (n=5); safe sex intention/attending HIV training program (n=5); HIV/STI/FP communication (n=4); anal/dry sex (n=3); pregnancy history (n=3); difficulty accessing condom (n=3); partner travels/stays away (n=2); relationship problems (n=1); hormonal contraceptive use (n=1); reason for condom use (n=1); circumcision (n=1) | Drug/alcohol use(n=20); sexual/physical violence/abuse (n=9); watching porn (n=6); sex debut (n=5); peer pressure/friends with sex experience (n=5); attending night club (n=4); communicating about sex/STI (n=3); having boy/girlfriend/sexually active (n=3); partner age/travels and reasons for travel (n=2); transactional sex (n=2); Condom less sex (n=2); pregnancy (n=2); rationale for partner choice (n=1); dry sex (n=1); relationship problems (n=1); meal preference (n=1); fighting/vandalism/theft/gambling (n=1); circumcision (n=1); internet/fashion use/knowing a foreigner (n=1) | Drug/alcohol use(n=6); having boy/girlfriend/sexually active (n=3); sexual/physical violence/abuse (n=2); watching porn (n=2); attending night club (n=2); discussing sexuality (n=1); relationship problems (n=1); attending a skills training program (n=1); internet use/knowing a foreigner (1); concurrency (n=1) | Drug/alcohol use(n=4); contraceptive/condom use (n=4); partner age difference/gender (n=3); category of 1st partner (n=2); sexual/physical violence/abuse (n=2); media access (n=1); motive of 1st sex (n=1); sex type/venue/anal sex position (n=1); using lubricant during sex (n=1); transactional sex (n=1); number of partners (n=1); relationship control (n=1); length of relationship (n=1) | Sex debut (n=3); having girlfriend/sexually active/sex frequency (n=3); relationship length (n=2); number of partners (n=2); alcohol/drug use (n=1); sexual/physical violence/abuse/IPV (n=1); circumcision status (n=1); partner age/age disparity (n=1); gestation at 1st ANC (n=1); intended pregnancy (n=1); travel/migration/partner lives in different village (n=1); seriousness of relationship (n=1); other venues attended/meeting sex partners at a venue (n=1); condom use (n=1); previous pregnancy/1st time mother (n=1) | Alcohol/drug use (n=1); peer influence (n=1); in a relationship (n=1); attending a skills training program (n=1) | Alcohol/drug use (n=5); watching porn (n=3); having girlfriend/sexually active (n=3); unexpected pregnancy/pregnancy/fathering a child (n=2); circumcision status (n=2); disco/night club attendance (n=2); travel out of city (n=1); partner age (n=1); relationship problems (n=1); peer pressure (n=1); technology use (n=1); condom access (n=1); sexting (n=1) |

| | | | | | | | |
|---|---|---|---|---|---|---|---|
| Knowledge, attitude & belief factors | knowledge of HIV status (n=16); HIV knowledge (n=11); perceived HIV/STI risk (n=6); attitude/perception towards condom use/self-efficacy (n=4); knowledge of someone died of HIV or HIV+ (n=3); HIV risk reduction self-efficacy (n=3); gender role perception/attitude/gender equity (n=3); perceived pregnancy risk/contraception attitude (n=2); attitude towards HIV+ people/HIV info (n=2); attitude around sexual control (n=2); HIV transmission beliefs (n=2); sex work stigma (n=1); IPV perception (n=1) | knowledge of HIV status (n=7); HIV/STI knowledge (n=2); perceived relationship/condom use self-efficacy (n=2); gender role perception (n=1); attitude around sexual control (n=1); knowledge of pregnancy prevention (n=1); perception of modern goods/fashion (n=1) | knowledge of HIV status (n=2); belief about reasons for having sex (n=1); beliefs about peer influence (n=1); self-efficacy/belief about one's ability to perform (n=1); knowledge of pregnancy prevention (n=1); attitude around sexual control (n=1); condom use self-efficacy (n=1); perception of modern goods/fashion (n=1); gender norms (n=1) | knowledge of HIV status/HIV testing (n=3); knowledge/perception of HIV risk (n=1); knowledge of pregnancy prevention (n=1); attitude around sexual control (n=1); condom use self-efficacy (n=1) | Knowledge of HIV status/tested for HIV (n=3); gender norms (n=2); gender role conflict (n=1) | Belief about reasons for engaging in sex (n=1); belief about peer influence (n=1); belief about one's ability to perform (n=1); knowledge of pregnancy prevention (n=1); attitude around sexual control (n=1); condom self-efficacy (n=1); knowledge of HIV status/HIV tested (n=1) | Perception/attention of HIV risk (n=3); knowledge of HIV status (n=2); knowledge of HIV/HIV prevention (n=3); source of HIV information (n=2); knowledge of FP (n=1); attitude towards HIV+ people (n=1); perceived risk of getting pregnant/making a girl pregnant (n=1); condom use attitude (n=1); sexual norms of social network (n=1) |
| Family & community factors | Family/community connectedness/social network affiliation (n=10); household income/wealth (n=7); parental residence/living arrangement (n=6); household education/father education/household education support (n=5); orphan status/loss of family member/ill family member (n=5); household occupants/size/family type (n=4); injury/disaster/accidents/war (n=2); uncertainty about the future/sense of future (n=2); caregiver of child (n=2); orientation towards success (n=1); trust in others/social participation (n=1); voluntary work (n=1); legal problems/crime (n=1); age of household head (n=1) | Household income/wealth (n=11); family/community connectedness/support/parental control/social network affiliation (n=9); parental residence/living arrangement (n=8); household occupants/size/family type (n=3); parents' marital status (n=2); household education (n=2); orphan status/loss of family member/ill family member (n=2); trust in others/social participation (n=1); legal problems/crime/injury (n=1); age of household head (n=1); voluntary work (n=1) | Parental residence/living arrangement (n=2); family connectedness/support /receiving money from parents/monitoring (n=2); legal problems/crime/injury (n=1); orphan status/loss of family member/ill family member (n=1); household size (n=1); household education (n=1); orientation towards success (n=1); uncertainty of the future (n=1); care giver of child (n=1); age of household head (n=1); household SES (n=1); closeness with camp network members/popularity/age of ego network (n=1) | Household SES/parental occupation (n=4); living arrangement (n=3); family/community/parental connectedness/concern/community group affiliation/support/peer relation/social network affiliation (n=3); having moved from an area (n=2); have children/number of children in household (n=2); parents' education (n=1); age of household head (n=1); orphan status (n=1); sense of future (n=1) | Social network affiliation/closeness of camp members (n=2); living arrangement (n=2); quality of parent/child relationship (n=1); parents' knowledge of child's out of home activities (n=1); parents'/child communication on sex issues (n=1); parents' SES (n=1); parents marital (n=1); camp members view concurrency as normative behavior (n=1); ego network popularity (n=1); age of ego network (n=1) | Parental connection/monitoring/relationship with parents (n=2); living arrangement (n=1); belonging to a group (n=1); voluntary work (n=1); age of household head (n=1); orientation towards success (n=1); uncertainty of the future (n=1); caregiver of child (n=1) | Living arrangement (n=6); parents' education (n=4); loss of family member/ill family member/orphan status (n=2); social network size/tie strength/homogeneity (n=1); family connectedness/support (n=1); legal problems/injury/crime (n=1); relationship to household head (n=1); duration of living in streets (n=1) |
| Mental/physical health factors | STIs (n=9); Depression (n=6); PTSD (n=3); bad health/recent sickness (n=2); self-esteem (n=2); ARV use (n=2); anxiety (n=1); psychosis (n=1); suicide ideation (n=1); number of VCT counselors/type of facility (n=1) | STIs (n=6); Depression (n=6); PTSD (n=3); bad health(n=1); ARV use (n=1); anxiety (n=1); psychosis (n=1); suicide ideation (n=1); disability (n=1) | STIs (n=1); Depression (n=1); bad health(n=1); suicide ideation (n=1) | STIs (n=1); suicide ideation (n=1); self-esteem (n=1) | STI (n=1) | Suicide ideation (n=1); STI (n=1) | Bad health (n=1) |
